# Genome-Wide Variations of End Motif in Cell-Free DNA Fragments Distinguish Immunotherapy Responders from Non-Responders in Head and Neck Cancer: A Multi-Institute Prospective Study

**DOI:** 10.64898/2026.03.24.26348354

**Authors:** Ravi Bandaru, Hailu Fu, Haizi Zheng, Jocelyn Liang, Li Wang, Shuchi Gulati, Benjamin H Hinrichs, Mingxiang Teng, Bin Zhang, Marsha Kocherginsky, Dechen Lin, David A. Hildeman, Francis P Worden, Matthew O Old, Neal E Dunlap, John M Kaczmar, Maura Gillison, Dalia El-Gamal, Trisha Wise-Draper, Yaping Liu

## Abstract

Reliable, minimally invasive biomarkers for predicting immunotherapy response in head and neck squamous cell carcinoma (HNSCC) remain an unmet clinical need. Here, using patients from a prospective, multi-institutional phase II clinical trial (NCT02641093), we performed whole-genome sequencing of 185 plasma cell-free DNA (cfDNA) samples collected longitudinally from 68 patients with locally advanced, surgically resectable HNSCC undergoing neoadjuvant and adjuvant pembrolizumab treatment. We developed the regional motif diversity score (rMDS), a novel fragmentomic metric quantifying the entropy of cfDNA 5′ end motifs across genomic regions. Remarkably, unsupervised analysis revealed that rMDS robustly distinguished immunotherapy responders from non-responders, outperforming established cfDNA fragmentomic metrics and copy number alterations, while demonstrating independence from technical confounders. Longitudinal analysis revealed dynamic rMDS changes in genomic regions enriched for immune-, lectin-, and keratinization-related genes–hallmarks of squamous cell carcinoma–reflecting the interplay between tumor and peripheral immunity during the immunotherapy treatment. Interestingly, the regions with the most dynamic rMDS changes were highly enriched in telomere-proximal loci, suggesting a novel link between telomere biology and cfDNA fragmentation. A machine learning classifier based on rMDS achieved robust predictive performance across multiple validation settings (AUC 0.89-0.99), with the highest accuracy at post-treatment timepoints and superior to PD-L1 expression and tumor fraction in the same sample. Predicted responders demonstrated significant trends toward improved disease-free survival (log rank test p=0.035, hazard ratio: 2.67, 95% confidence interval: 1.03-6.92), underscoring the clinical utility of rMDS-based stratification. These findings position rMDS as a biologically meaningful and clinically actionable biomarker for immunotherapy response in HNSCC, supporting its integration into future risk assessment frameworks and broader cancer care.

## Introduction

Head and neck squamous cell carcinoma (HNSCC) is the seventh most prevalent cancer globally, characterized by high recurrence rates (∼50%) and poor survival despite aggressive multimodal treatment strategies^1–3^. Immune checkpoint blockers (ICBs), particularly those targeting programmed death-(ligand)-1 (PD-(L)1), such as pembrolizumab and nivolumab, have demonstrated promising clinical outcomes, significantly benefiting select patient groups^4,5^. However, primary resistance remains prevalent, with only approximately 18% of patients with HNSCC achieving substantial responses to immunotherapy^5,6^. This highlights an urgent need for reliable biomarkers capable of predicting immunotherapy responses to facilitate personalized therapeutic strategies. Current biomarkers, including PD-L1 expression, tumor mutational burden, and immune gene expression profiles, have been extensively investigated but demonstrate limited sensitivity and specificity in differentiating responders from non-responders and frequently require invasive tissue biopsies^7^.

Fragmentation patterns of circulating cell-free DNA (cfDNA) in plasma have recently emerged as a promising class of minimally invasive biomarkers for predicting and monitoring therapeutic response in cancer, particularly in the context of immunotherapy^8–10^. cfDNA, which is released into the bloodstream through cellular apoptosis, necrosis, and active secretion, carries a wealth of information reflecting the genomic and epigenomic landscape of its cells of origin^11–16^. Recent advances in sequencing technologies and bioinformatics have enabled detailed analysis of cfDNA fragmentation patterns, such as fragment size distributions, genome-wide coverage, DNA evaluation of fragments for early interception (DELFI)^17^, and the sequence context of fragment ends^18^. Particularly, end motifs of cfDNA fragments and derived metrics like motif diversity score (MDS) have emerged as robust fragmentomic biomarkers^18^. For example, characteristic motif alterations at cfDNA fragment ends and elevated MDS have been consistently observed in HNSCC and other malignancies^18^. These fragmentomic features have demonstrated potential to detect the presence of cancer, infer tissue of origin, and track disease progression with high sensitivity and specificity^11,12^.

Building on these advances, several studies have sought to explore the clinical utility of cfDNA fragmentation patterns as biomarkers for immunotherapy response in cancer patients^8,9^. However, these investigations have been retrospective, and single-institution efforts. cfDNA fragmentation was analyzed primarily as a supplementary metric to enhance the detection of copy number alterations, rather than as a standalone biomarker. Moreover, these studies only included a small number of patients per cancer type or pooled samples across diverse malignancies, limiting the generalizability and interpretability of their findings. Notably, to date, no prospective studies have systematically evaluated cfDNA fragmentation features as predictive or prognostic biomarkers for cancer immunotherapy across multiple institutes, especially in patients with HNSCC.

In a recent multi-institutional prospective phase II clinical trial (NCT02641093)^19^, we investigated the efficacy of neoadjuvant and adjuvant pembrolizumab in patients with surgically resectable, locally advanced HNSCC^19^. The trial demonstrated improved one-year disease-free survival (DFS) in intermediate-risk patients compared to historical controls. These findings were recently validated by phase III KEYNOTE-689 (NCT03765918), which showed improved disease-free survival with neoadjuvant and adjuvant pembrolizumab and established this approach as a new standard of care for locally advanced HNSCC^20^. Nonetheless, conventional predictive biomarkers, including PD-L1 expression and immune gene signatures, proved inadequate in accurately stratifying patient responses, highlighting the limitations of existing molecular predictors. Motivated by these findings, we explored whether cfDNA fragmentation patterns could predict response to pembrolizumab immunotherapy in patients enrolled in this clinical trial. We conducted longitudinal analyses using low-coverage (∼1.9x) cfDNA whole-genome sequencing (WGS) of plasma samples collected at three different time points pre- and post-treatment, ensuring balanced demographic characteristics between responder and non-responder groups. We developed a novel fragmentomic metric, termed regional motif diversity score (rMDS), designed to quantify genome-wide variations in the entropy of end motifs across distinct genomic regions. Remarkably, unsupervised analysis using rMDS effectively distinguished immunotherapy responders from non-responders and outperformed conventional fragmentomic features and other biomarkers from the same patient cohort. This approach captures clinically significant changes in cfDNA fragmentation undetectable by traditional biomarkers, thereby providing a non-invasive method for response monitoring and enhancing clinical risk stratification.

## Results

### Generation of cfDNA WGS from longitudinal plasma samples in a multi-institutional Phase II trial

Our cohort comprises 68 patients from the original cohort of 92 patients with locally advanced, surgically resectable HNSCC who have available plasma and are enrolled in a prospective, multi-institutional phase II clinical trial. For a subset of patients in the original cohort, only whole blood was collected, and these samples were excluded because genomic DNA contamination precludes accurate cfDNA fragmentation profiling. Thus, the current cohort spanned six academic institutes and included 40 clinical full or partial responders and 28 non-responders, as determined by treatment effect **(see Methods)**, with no significant differences in demographic or clinical variables between response groups **(Table 1**, **Fig. 1, Supplementary Table 1)**. All patients received a single 200 mg intravenous (IV) dose of neoadjuvant pembrolizumab, followed by surgical resection and adjuvant radiotherapy (60–66 gray [Gy]). High-risk patients received concurrent weekly cisplatin (40 mg/m^2^), and all patients were eligible to receive post-operative pembrolizumab for up to seven total doses. 0.5 ml of plasma was collected at three defined clinical timepoints: prior to neoadjuvant therapy (Screen), the day of surgery (Day 0), and 3-10 weeks post-surgery (Adjuvant Week 1).

**Table 1.**
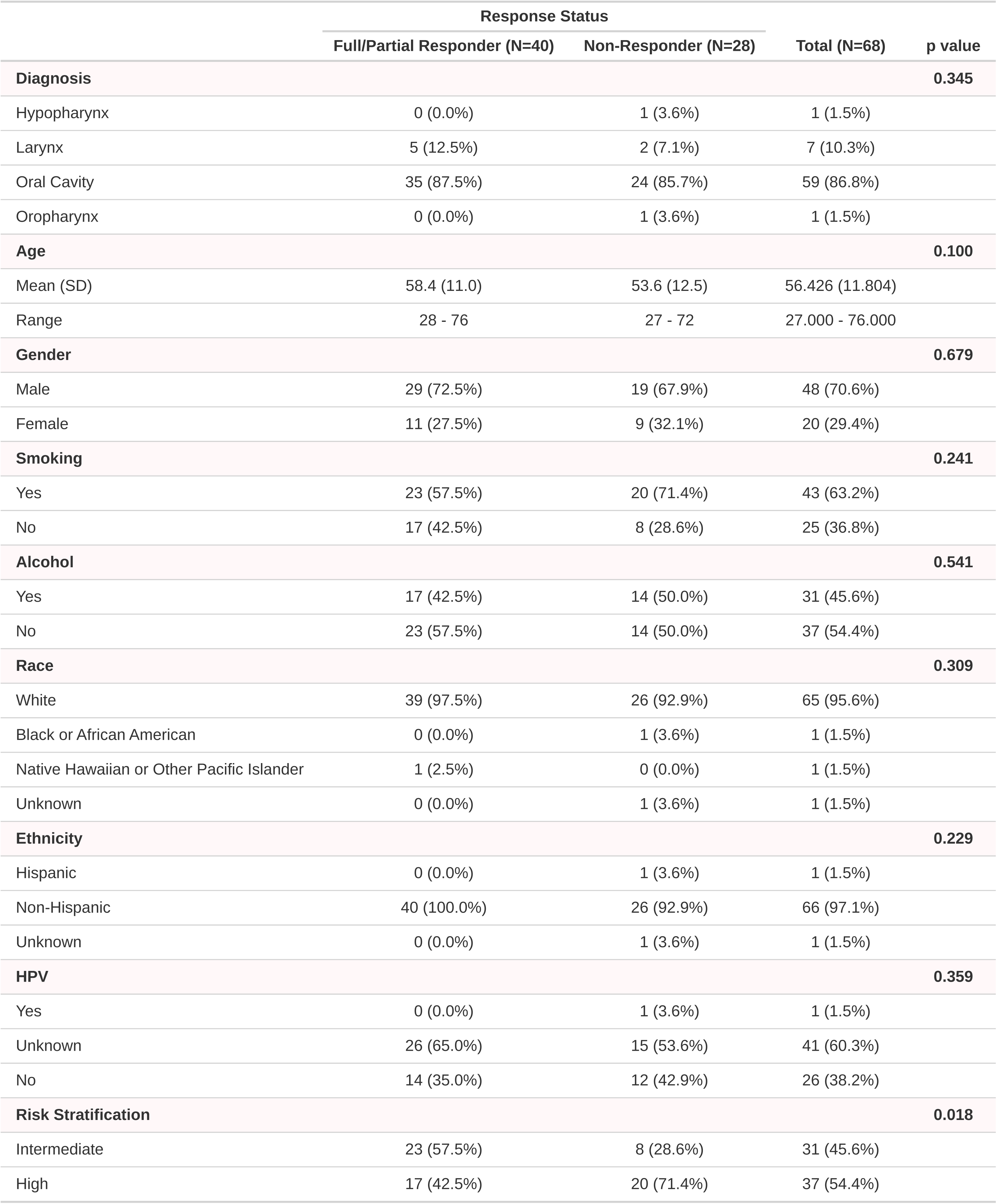
Metadata of the study cohort.

**Figure 1.**
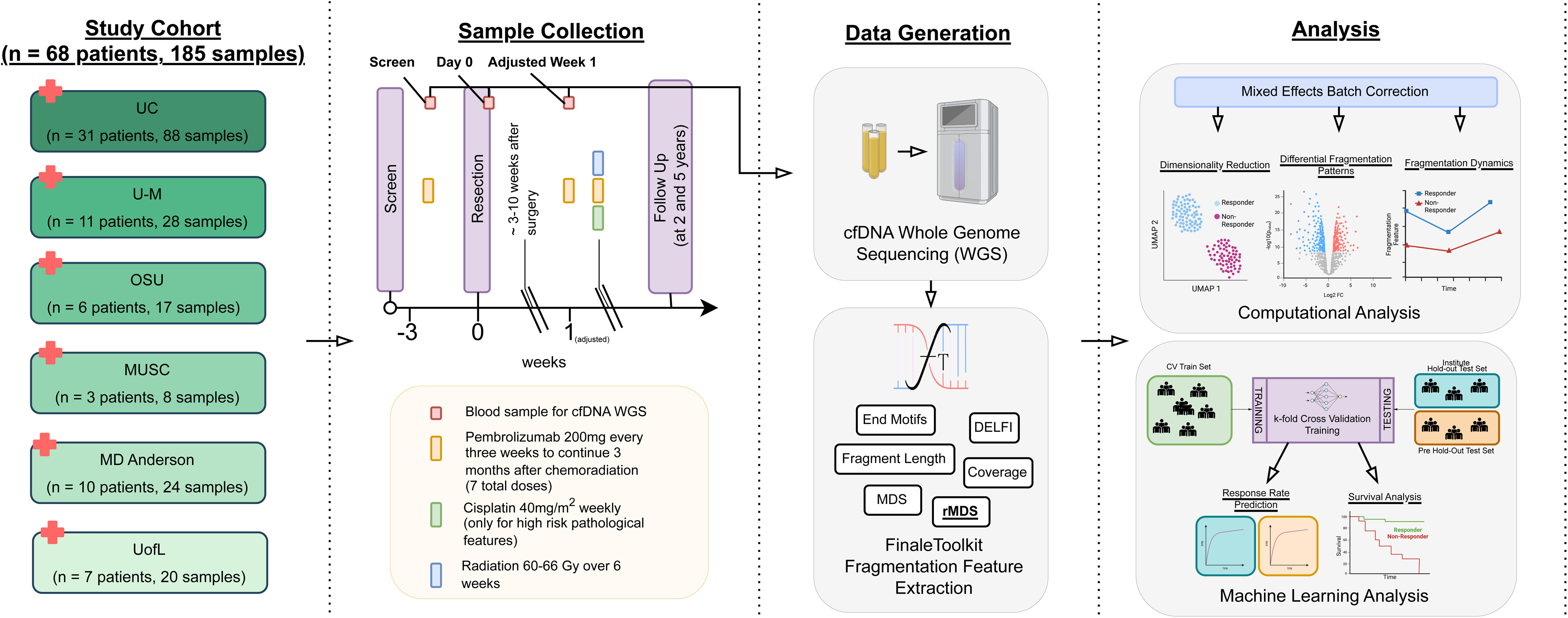
Experimental design and analytical workflow for the HNSCC cohort. Patients with head and neck squamous cell carcinoma (HNSCC) underwent surgical tumor resection and Pembrolizumab immunotherapy, followed by adjuvant radiation therapy (60–66 Gray [Gy] over 6 weeks), and cisplatin (40mg/m3) for high-risk pathological features. 185 blood samples were collected at defined intervals throughout the course of treatment and follow-up from 68 patients across six institutions: University of Cincinnati (UC), University of Michigan (U-M), Ohio State University (OSU), Medical University of South Carolina (MUSC), The University of Texas MD Anderson Cancer Center (MDACC), and the University of Louisville (UofL). Plasma-derived circulating cell-free DNA (cfDNA) was isolated from blood samples collected at three different time points (Screen, Day 0, and Adjuvant Week 1) and subjected to whole-genome sequencing (WGS). Fragmentation features were extracted using *FinaleToolkit,* including Motif Diversity Score (MDS, single genome-wide summary score), regional Motif Diversity Score (rMDS), end motif frequencies, DNA Evaluation of Fragments for Early Interception (DELFI), genome-wide coverage, and fragment length distributions.

Of the 185 cfDNA WGS libraries generated from plasma samples, 176 met predefined quality control criteria (details provided in Methods), achieving a median sequencing coverage of approximately 1.9x per sample (**Supplementary Fig. 1A, Supplementary Table 2**). From these, 10 patients (i.e., 25 samples) were randomly set aside as a pre-specified holdout set for the later evaluation of the machine learning model, and the analysts remained blinded to these samples. The remaining 151 samples constituted the analysis dataset used for all subsequent analyses. (**Supplementary Fig. 1B**). Forty-one patients provided samples across all three longitudinal time points, with sample distribution approximately balanced among visits (**Supplementary Fig. 2A**). Samples from the University of Cincinnati (UC) constituted a greater proportion (48%) compared to other institutions (**Supplementary Fig. 2B**). Responders and non-responders were balanced in the analysis set.

Technical metadata, including cfDNA isolation and library preparation dates, showed no systematic biases or imbalances between analysis set and pre-holdout set (**Supplementary Fig. 2C-D**).

### Unsupervised analysis of regional Motif Diversity Score (rMDS) distinguishes immunotherapy responders from non-responders

We first examined whether differences in fragmentation patterns existed between responders and non-responders. Given the low sequencing coverage, we partitioned the reference genome into non-overlapping bins of 500 kb. Within each genomic bin, we utilized FinaleToolkit^21^ to extract standard fragmentomic features, including fragment length, fragment coverage corrected for GC-content bias, and DELFI. Additionally, FinaleToolkit was used to compute genome-wide summary statistics for each sample, including frequencies of 5′ end motifs (n=256) and MDS (n=1)^18^. Copy number variations (CNVs) for each genomic bin and tumor fractions for each sample were calculated using ichorCNA^22^. To isolate response-associated variance, we removed potential batch effects arising from plasma collection sites, cfDNA isolation procedures, and library preparation dates (details provided in Methods; **Supplementary Fig. 3**). Despite these analyses, none of these features demonstrated the separations between responders and non-responders (silhouette scores 0.015-0.077, **Supplementary Fig. 4, Supplementary Table 3**).

A recent study has reported differences in genome-wide MDS between HNSCC patients and healthy individuals^18^. However, genome-wide MDS represents only a single aggregated metric of the variations in cfDNA 5′ end motifs. Given that the 5′ end of cfDNA fragments strongly correlates with cellular epigenetic profiles^18,23^, which vary significantly across genomic regions and reflect underlying gene-regulatory mechanisms^24,25^, evaluating end motif diversity variations at a regional level across the genome could better capture this biological complexity. To address this, we developed a novel fragmentomic metric, termed the regional motif diversity score (rMDS), which extends genome-wide MDS by quantifying variations in the Shannon entropy of 5′ end 4-mer motifs across distinct genomic regions **(Fig. 2A)**. Remarkably, after correcting for batch effects, the z-score transformed rMDS value effectively and unbiasedly discriminated immunotherapy responders from non-responders in our cohort **(Fig. 2B, left)**. Interestingly, rMDS could differentiate between these two groups even at the pre-treatment screening timepoint, suggesting that baseline differences in cellular epigenetic states may predict responsiveness to immunotherapy **(Fig. 2C)**. Greatest separation was observed at the most recent sample collection timepoint (Adjuvant Week 1, silhouette score=0.327), compared to Day 0 (silhouette score=0.297) and Screening (silhouette score=0.261).

**Figure 2.**
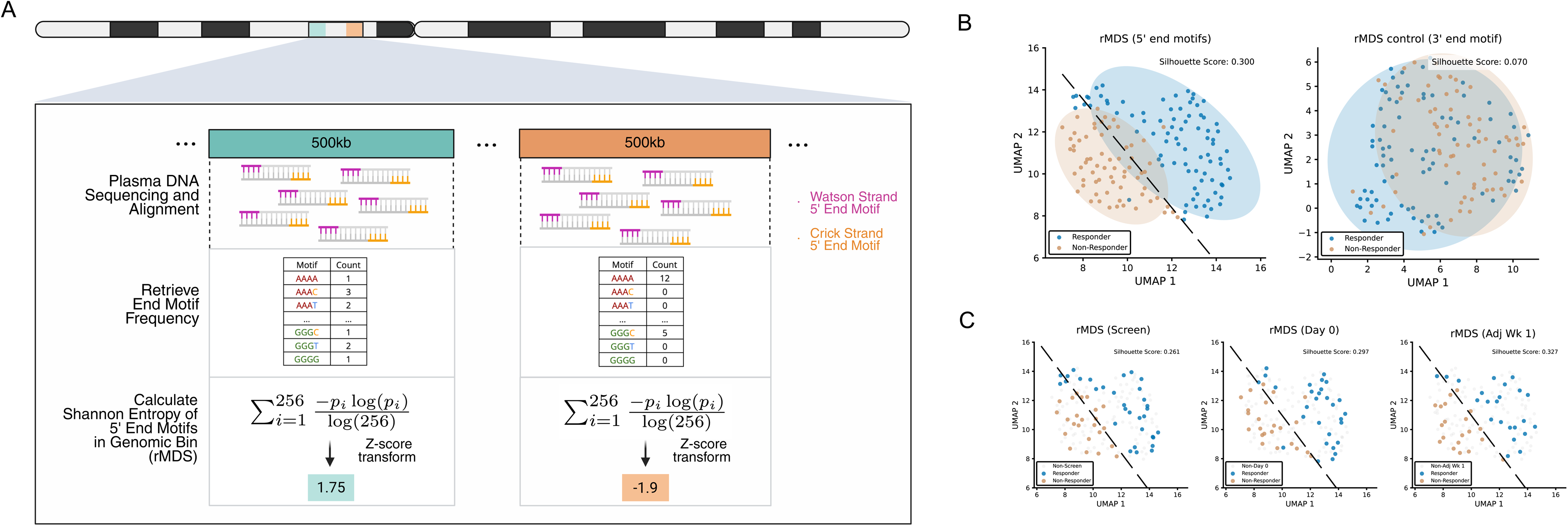
Regional motif diversity score (rMDS) distinguishes immunotherapy responders from non-responders. **(A)** Schematic representation of rMDS calculation: cfDNA 5′ end motifs are quantified within 500 kb genomic bins, and Shannon entropy is computed to yield rMDS, preserving regional information relative to genome-wide MDS. **(B)** UMAP visualization demonstrates clear separation of responders and non-responders based on rMDS from 5′ end motifs that are related to cfDNA fragmentation, with minimal separation observed using 3′ end motifs as the control. **(C)** rMDS (5′ end motifs) profiles at Screening, Day 0, and Adjuvant Week 1 also independently separate responders from non-responders. Ellipses represent ∼2 standard deviations from group centroids.

It is possible that the observed separation between the two groups could be attributed to potential batch effects specifically related to nucleotide sequence context or confounding changes in the overall nucleotide composition of cfDNA fragments, rather than genuine fragmentation patterns at the 5′ end. To address these concerns, we calculated rMDS control based on 3′ end 4-mer motifs, which are not reported to be related to cfDNA fragmentation by the standard WGS approach we used. As expected, we found no differences in 3′ end rMDS control between responders and non-responders (silhouette score=0.07), suggesting that the observed group separation by 5′ end rMDS is not confounded by sequence composition or other technical effects (**Fig. 2B, right).**

To assess the impact of sequencing depth on rMDS estimation, we performed empirical downsampling using the deeply sequenced BH01 cfDNA dataset from Snyder et al.^13^ (∼56x effective coverage). Reads were downsampled to 50x, 25x, 10x, 5x, 2x, 1x, and 0.1x coverage, and 500 kb rMDS profiles were recalculated **(Supplementary Fig. 6).** Correlation with the full-depth reference increased monotonically with coverage, reaching r = 0.93 at 5x and remaining high at ∼2x (r = 0.82), comparable to our study (∼1.9x). Substantial decreases in correlation with the full-depth reference occurred only at extreme downsampling (0.1x). Downsampling of patient samples similarly demonstrated preserved responder/non-responder discrimination at moderate reductions in coverage (silhouette scores 0.3-0.42), with deterioration only at very low depth (silhouette score=0.151; **Supplementary Fig. 7).** These results suggest that while deeper sequencing improves precision, biologically meaningful rMDS structure is retained at the coverage used in our study.

We further evaluated the optimal k-mer choice **(Supplementary Fig. 8)**. The 4-mer model demonstrated the strongest group separation (silhouette score=0.3). Shorter 3-mers likely lack sufficient sequence complexity, whereas 5- and 6-mers greatly expand the feature space (1,024-4,096 different end motifs), resulting in sparse counts per end motif type in each genomic bin at our sequencing depth (∼1-4 fragments per end motif type in each bin) and, thus, unstable entropy estimation. Therefore, 4-mers provide an optimal balance between contextual information and statistical robustness under the current low-coverage conditions.

### Longitudinal dynamics of differential rMDS patterns reveal telomere-associated fragmentation shifts

To identify genomic regions with differential rMDS patterns, we conducted a genome-wide comparative analysis of rMDS between immunotherapy responders and non-responders. Besides the technical batch effect, other clinical variables, such as age and gender, could also affect the cfDNA fragmentation status. We further employed a linear mixed-effects modeling approach to account for repeated measures across timepoints and different clinical covariates, including age, race, gender, ethnicity, diagnosis, smoking status, and alcohol use (details in Methods). After the correction for multiple hypothesis testing, we identified 1,080 significantly differential 500 kb bins (q value < 0.1, **Supplementary Table 4**), including 545 genomic bins exhibiting increased rMDS in non-responders and 535 bins showing decreased rMDS (**Fig. 3A**). To explore the longitudinal dynamics of these differential rMDS regions, we applied joint hierarchical clustering to z-score transformed rMDS values measured across three longitudinal time points in the same patients (**Fig. 3B, top panel**).

**Figure 3.**
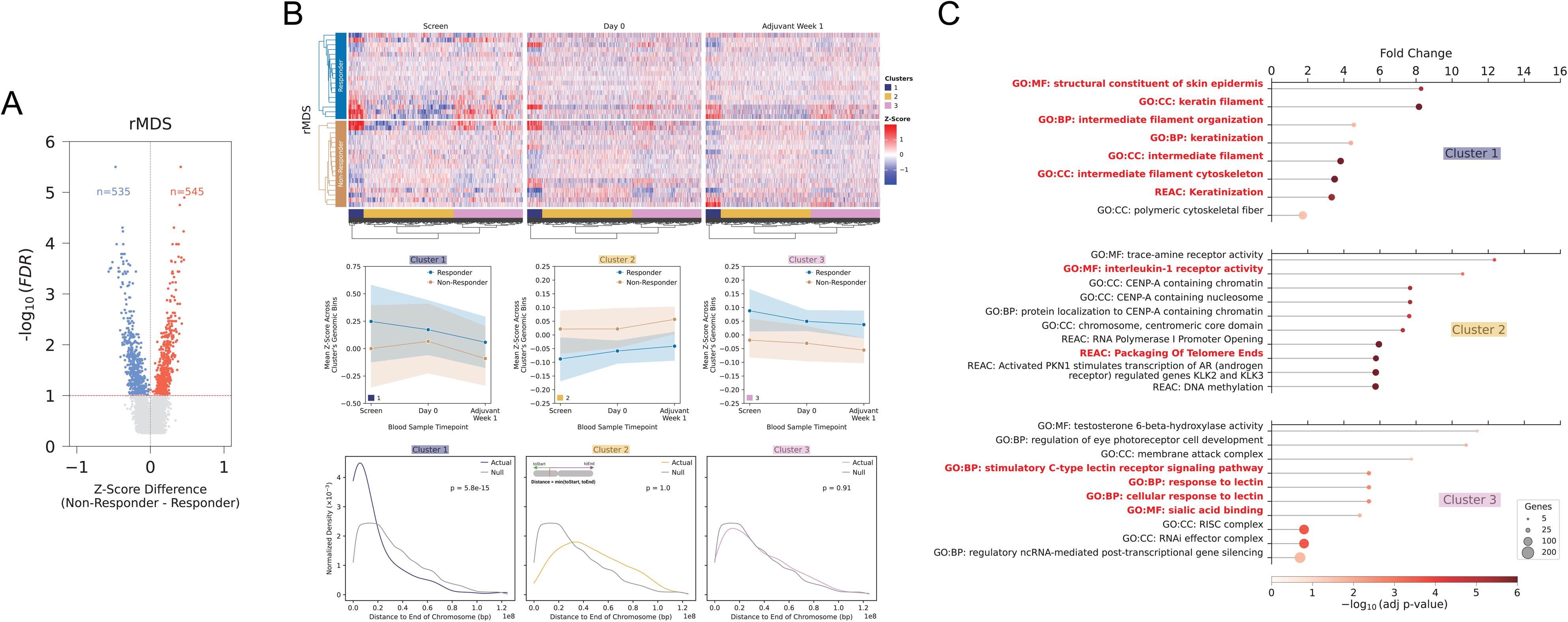
Differential rMDS regions between immunotherapy responders and non-responders. **(A)** Volcano plot of differential rMDS genomic bins between responders and non-responders across all samples. Regions with significantly increased or decreased motif diversity in responders are highlighted (FDR[<[0.1). **(B)** Heatmap of differentially enriched rMDS regions across three timepoints (Screen, Day 0, Adjuvant Week 1) reveals distinct longitudinal dynamics between responders and non-responders, stratified by cluster. Middle panel, line plots show the mean z-score trajectory over time for each cluster, separated by response status. Bottom panel shows the normalized density of distances from each rMDS region to the nearest chromosome end for each cluster, compared to a null distribution; p-values indicate one-sided Mann-Whitney U tests assessing whether the actual distribution is significantly closer to chromosome ends than the null. **(C)** Gene ontology enrichment analysis of differential rMDS clusters.

This clustering approach yielded three distinct clusters, each characterized by unique dynamic patterns of rMDS. Cluster 1 showed the most pronounced divergence in rMDS between responders and non-responders, with initially large differences at screening that narrowed at Day 0 before separating again by Adjuvant Week 1. Cluster 2 consistently showed higher rMDS in non-responders across all time points, whereas Cluster 3 demonstrated the opposite pattern (**Fig. 3B middle panel**).

Because the differential regions were defined using a statistical model that jointly evaluated response-associated differences across all timepoints while adjusting for patient-level covariates, we further examined the trajectories using covariate-corrected rMDS values. Consistent with the model results, visualization of covariate-adjusted signals confirmed significant differences between responders and non-responders for all three clusters (Supplementary Fig. 9), supporting the robustness of the identified differential regions. These analyses clarify that the regions identified by the mixed-effects model capture longitudinal response-associated effects that may not be fully apparent when visualizing unadjusted, only technically-corrected, rMDS values alone.

Intrigued by the genomic locations of these clusters, we performed enrichment analysis. Remarkably, Cluster 1 regions were significantly enriched near telomeric regions (Mann-Whitney p = 5.8×10^−15^, permutation p < 1×10^-5^), a feature absent in Clusters 2 and 3 (**Fig. 3B, bottom panel**). These findings suggest that distinct fragmentation dynamics at telomeric chromatin may be associated with the immunotherapy response.

### Gene ontology analysis links longitudinal dynamics of differential rMDS patterns to immune, lectin signaling, and keratinization programs

Next, we sought to characterize the genes enriched in differential rMDS regions exhibiting distinct longitudinal dynamics. Gene ontology (GO) analysis identified unique functional signatures across rMDS clusters related to both immune system components and HNSCC tumor biology **(Fig. 3C, Supplementary Table 5)**. Specifically, Cluster 1 was characterized by enrichment in keratinization, a defining histologic hallmark of squamous cell carcinomas, which constitute the predominant form of head and neck cancers. Particularly, the extent and type of keratinization was known to have prognostic value and were linked to differential survival outcomes^26–28.29–32^. Cluster 2 was predominantly enriched for Interleukin-1 (IL-1) and Centromere protein A (CENP-A)-related terms. Previous studies indicate that IL-1 signaling has been related to tumor growth and metastasis in HNSCC^33^. Moreover, subnuclear patterns of CENP-A have been shown to be correlated with immunotherapy responses in other cancer types^34^ and chemotherapy disease curability in HNSCC patients^35^. Cluster 3 was characterized by enrichment in lectin signaling; previous studies have shown that genes within this group, such as KLRK1, are highly expressed in HNSCC and are linked to favorable prognosis and increased immune cell infiltration^36^.

Representative rMDS Z-score trajectories at selected genomic loci further illustrated these patterns. A Cluster 3 region located at chr12:52.0–52.5 Mb, encompassing keratin genes (e.g., KRT7, KRT86, KRT85), showed an increase in rMDS in responders and a decrease in rMDS in non-responders **(Supplementary Fig. 10)**. Another telomeric Cluster 1 region at chr9:135.5–137.5 Mb demonstrated similar diverging trends **(Supplementary Fig. 10).** Furthermore, loci enriched for CENP-A-related and lectin signaling-related genes from Clusters 2 and 3 displayed distinct group-specific fragmentation dynamics **(Supplementary Fig. 11)**.

### rMDS-based predictive modeling robustly discriminates immunotherapy responders across multiple validation scenarios

We next explored the potential of rMDS as a composite biomarker for predicting immunotherapy response in HNSCC patients. To mitigate overfitting due to limited sample size, we applied singular value decomposition (SVD) to genome-wide rMDS values, reducing feature dimensionality. Additionally, we utilized the Tabular Prior-data Fitted Network (TabPFN) classifier, specifically designed for machine learning studies with small sample sizes^37^. Patient-level predictions were obtained by using the most recent available time point due to our unsupervised analysis suggesting greater rMDS separation at more recent timepoints (**Fig. 2C, 4A**).

**Figure 4.**
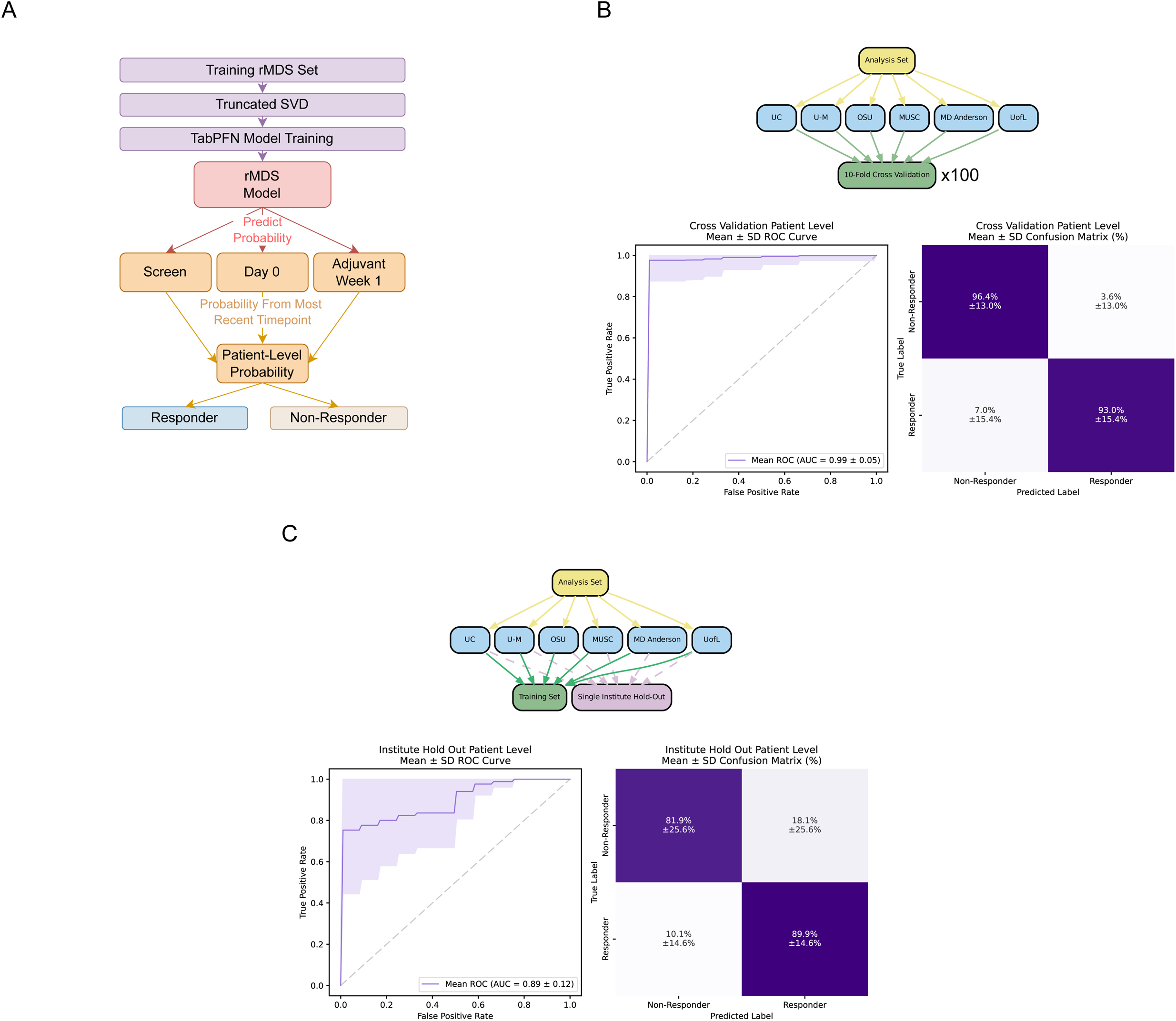
Predicting immunotherapy response in HNSCC using rMDS. **(A)** Overview of the rMDS-based machine learning pipeline. Dimensionality reduction was first applied using truncated singular value decomposition (SVD). Dimensionality reduction was performed by truncated SVD, and reduced features were used to train TabPFN models. Patient-level probabilities were obtained by using the most recent available time point, then binarized as Responder or Non-Responder. **(B)** Cross-validation strategy. Models were trained using a 10-fold patient-level cross-validation repeated 100 times. **(C)** Institute hold-out evaluation strategy. For each iteration, one institute was held out entirely for testing while the model was trained on the remaining institutions. The average ROC curve and confusion matrix (mean ± standard deviation) are shown.

We implemented a comprehensive three-tier validation strategy to rigorously evaluate model performance under different scenarios. First, we assessed model accuracy through 10-fold patient-level cross-validation repeated 100 times on the analysis set. This approach yielded exceptional performance with a mean AUC of 0.99 ± 0.05 **(Fig. 4B)**. At 95% specificity, the model achieved 97.6% ± 10.1% sensitivity. Classification accuracy was consistently high across response categories, correctly identifying non-responders 96.4% ± 13.0% of the time and responders 93.0% ± 15.4% of the time **(Fig. 4B)**. Sample-level analysis confirmed robust performance across all collection timepoints, with the highest AUC observed at Adjuvant Week 1 (0.99 ± 0.04), followed by Screen (0.97 ± 0.09) and Day 0 (0.95 ± 0.12) **(Supplementary Fig. 12, Supplementary Table 6)**.

To evaluate real-world applicability where samples originate from previously unseen institutions, we performed single-institute holdout validation within the analysis set (leave-one-institute-out; LOIO). Each institution was sequentially held out while training on samples from all other sites. This more stringent evaluation yielded a mean AUC of 0.89 ± 0.12 **(Fig. 4C)**, with a sensitivity of 75.3% ± 30.9% at 95% specificity. Classification accuracy remained robust, with correct identification of non-responders and responders 81.9% ± 25.6% and 89.9% ± 14.6% of the time, respectively **(Fig. 4C)**. Consistent with previous analyses, Adjuvant Week 1 samples showed the highest discriminatory power (AUC = 0.94 ± 0.14), followed by Screen (0.83 ± 0.15) and Day 0 (0.82 ± 0.21) **(Supplementary Fig. 13, Supplementary Table 7)**.

We assessed model generalizability by simulating the scenario of applying our predictor to entirely independent patient cohorts when the batch effects were not fully corrected. This pre-holdout analysis was included specifically to illustrate the impact of uncorrected technical variation on the predictive model. We randomly selected 10 patients (and their associated longitudinal samples) as a “pre-holdout” set across 100 iterations. This most stringent evaluation scenario yielded a mean AUC of 0.69 ± 0.19 **(Supplementary Fig. 14)**, with a sensitivity of 37.9% ± 27.0% at 95% specificity. While classification accuracy was more modest under these conditions, correctly identifying non-responders 57.2% ± 26.5% of the time and responders 68.5% ± 20.0% of the time, performance remained above chance levels across all timepoints (AUC range: 0.67-0.68) **(Supplementary Fig. 15, Supplementary Table 6)**. Together, these results demonstrate that when technical batch effects are properly accounted for, rMDS-based prediction models maintain robust performance under multiple validation conditions.

To further quantify the impact of batch correction on predictive performance, we directly compared model accuracy before and after correction in both cross-validation and institute hold-out settings **(Supplementary Fig. 16)**. In the absence of correction, performance was markedly reduced (cross-validation mean AUC: 0.51 ± 0.28; institute hold-out mean AUC: 0.64 ± 0.20). After correction, performance improved substantially (10-fold cross-validation mean AUC: 0.99 ± 0.05; institute hold-out mean AUC: 0.89 ± 0.12).

Given the enrichment of telomeric and keratinization-associated loci in differential rMDS clusters, we also evaluated whether restricting features to these focal regions retained predictive value (**Supplementary Fig. 17)**. Models trained using rMDS derived exclusively from keratinization loci showed moderate discrimination (mean AUC = 0.67 ± 0.21), whereas telomere-proximal regions demonstrated improved performance (mean AUC = 0.76 ± 0.28). Combining keratinization and telomeric loci maintained comparable performance (mean AUC = 0.73 ± 0.32). These results suggest that specific biologically enriched regions harbor concentrated fragmentation signals that may serve as feature subsets for future biomarker development with increased coverage.

### rMDS-based response predictions correlate with survival outcomes and enhance risk stratification

We next evaluated whether rMDS-based response predictions could serve as prognostic biomarkers. Five-year disease-free survival (DFS) and overall survival (OS) were analyzed based on predicted response status using the single-institute holdout evaluation. Notably, despite the single institute hold-out achieving 74.2% accuracy, patients predicted as responders exhibited significantly improved DFS compared to predicted non-responders (log-rank test, p = 0.035; **Fig. 5A**). This prognostic capability was superior to established biomarkers, as no significant differences were observed when using PD-L1 combined positive score (CPS) (p = 0.97) or tumor fraction (p = 0.50) from the same patients. The rMDS predictions demonstrated comparable prognostic value to pathological risk stratification (p=0.013; **Supplementary Fig. 18**).

**Figure 5.**
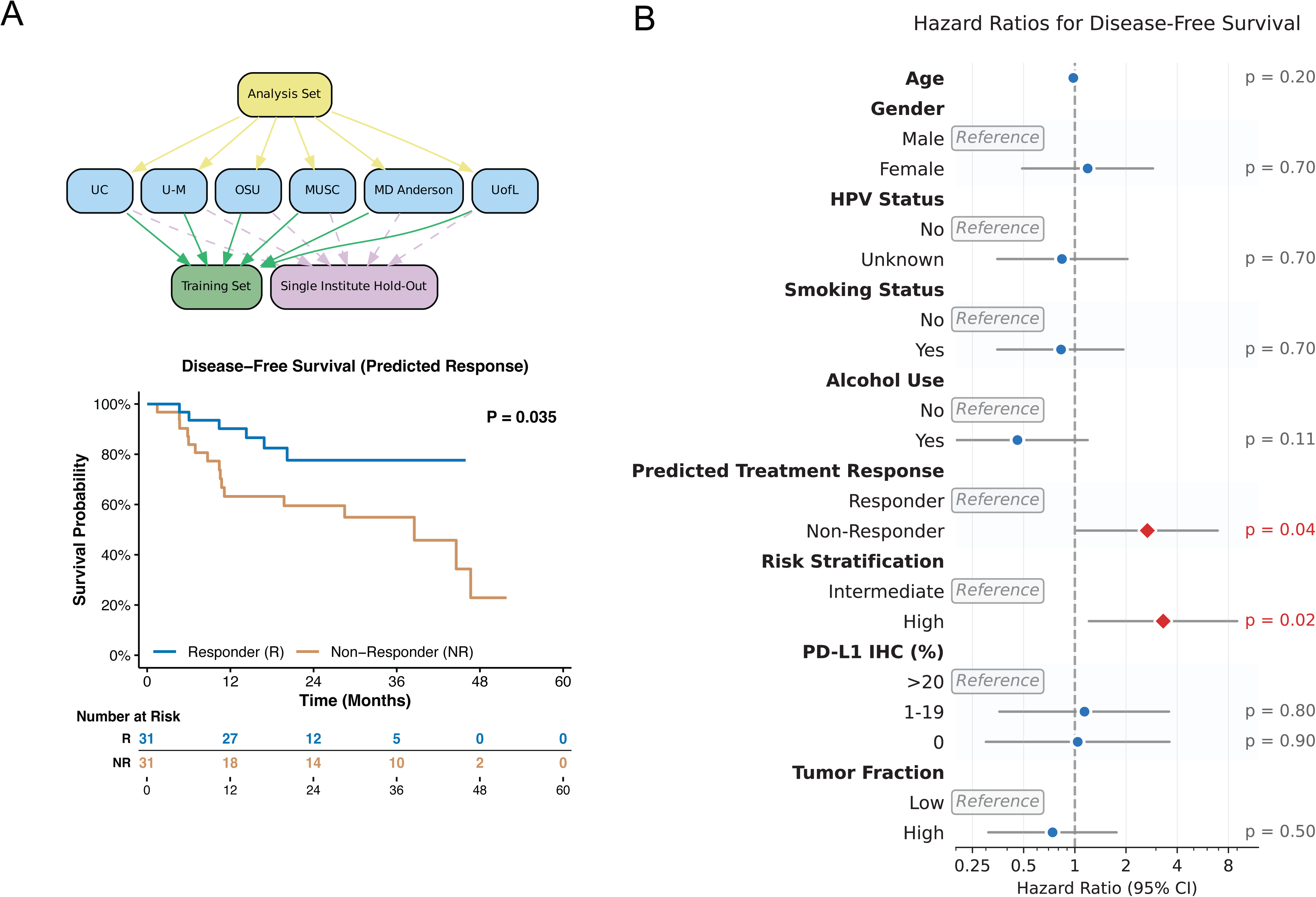
cfDNA rMDS-based model predicts clinical outcomes in pembrolizumab-treated HNSCC patients. **(A)** Disease-free survival stratified by predicted treatment response (Responder vs Non-Responder) using a cfDNA rMDS-based model with a single-institute hold-out evaluation strategy. The global log-rank p-value and risk table are shown. **(B)** Hazard ratios for disease-free survival across clinical and demographic covariates (p-values indicated).

For overall survival, while rMDS-predicted responders showed a trend toward improved outcomes (log-rank test, p = 0.197), this did not reach statistical significance. In comparison, neither PD-L1 CPS (p = 0.52) nor tumor fraction (p = 0.15) showed prognostic value for OS, whereas pathological risk remained significant (p=0.002, **Supplementary Fig. 19A-D**). Hazard ratio analysis revealed that rMDS-predicted non-responders had significant hazard ratios (HR) of 2.67 (95% confidence interval CI: 1.03-6.92) for DFS, demonstrating the clinical utility of this biomarker approach. While no statistically significant HR was observed for OS with rMDS predictions (1.98, 95% CI: 0.69-5.74), pathological risk showed significant HRs for both DFS (3.31, 95% CI: 1.21-9.01) and OS (7.16, 95% CI: 1.62-31.6) (**Fig. 5B, Supplementary Fig. 19E, Supplementary Table 7**). Since some patients may have experienced recurrence between neoadjuvant and adjuvant treatments, we further adjusted the survival time to reflect months since neoadjuvant pembrolizumab initiation, and the results remained similar. (**Supplementary Fig. 20-21, Supplementary Table 7).**

These findings suggest that rMDS-based predictions may provide clinically meaningful risk stratification and short-term outcome prediction, outperforming conventional markers like PD-L1 CPS and tumor fraction and offering the potential to enhance future clinical risk stratification models.

## Discussion

This study represents, to our knowledge, the largest prospective, multi-institutional investigation of cfDNA fragmentation in HNSCC to date. We demonstrate that the diversity of cfDNA end motifs, quantified by the regional Motif Diversity Score (rMDS), can distinguish immunotherapy responders from non-responders in an unsupervised manner. Utilizing low-coverage WGS data from a cohort treated with neoadjuvant pembrolizumab, we show that rMDS separates responders from non-responders with high accuracy and correlates with disease-free survival more robustly than conventional biomarkers, such as PD-L1 expression or tumor fraction.

Our study provides several novel insights. First, rMDS, derived from the entropy of 5′ end motifs in 500 kb bins, captures the longitudinal dynamics of region-specific fragmentation changes in an unsupervised manner, outperforming other fragmentomic features and genetic aberrations. Unlike previous fragmentomic approaches that summarized end-motif diversity across the entire genome, rMDS quantifies regional variation, capturing biologically localized chromatin differences associated with immunotherapy response. Remarkably, rMDS was able to distinguish responders from non-responders even at the pre-treatment screening timepoint, suggesting that baseline cellular epigenetic plasticity may predict future responsiveness to immunotherapy. This observation warrants further experimental validation using matched tumor and immune cell samples. While prior studies have shown the utility of genome-wide MDS in distinguishing HNSCC patients from healthy individuals^18^, our results highlight the limitations of relying on a single summary statistic for predicting treatment response. The original MDS compresses the diversity of 5′ end motifs into a single value and ignores the baseline variation across individuals, potentially masking important regional fragmentation dynamics.

Hierarchical clustering of differentially fragmented regions revealed distinct patterns of divergence between responders and non-responders, with region-specific enrichments originating from both tumor and immune compartments, suggesting that tumor-immune interactions shape patients’ responses to immunotherapy. Clusters 2 and 3 comprised loci with persistent baseline differences and were highly enriched for immune, CENP-A, and lectin signaling-related genes, which have previously implicated immunotherapy responses in other cancer types and chemotherapy treatment outcomes, as well as HNSCC disease progression and prognosis. In contrast, Cluster 1 contained regions exhibiting more dynamic changes following treatment and was enriched for keratinization-related genes—the histologic hallmark of squamous cell carcinoma—highlighting the potential role of tumor-derived cfDNA or alterations in tumor cell epigenomes during immunotherapy. Interestingly, Cluster 1 regions were preferentially localized near telomeric ends, which are highly associated with genomic instability^38^ and may reflect different cell death mechanisms, such as apoptosis and necrosis. Emerging evidence also suggests that telomere maintenance genes influence immune cell infiltration and response to immunotherapy^39,40^. However, the relationship between telomere biology and cfDNA fragmentation, particularly regarding end motif diversity, remains unexplored and needs further investigation.

Our study also has several limitations. The relatively low sequencing coverage (∼1.9x) and patient heterogeneity limited our ability to investigate rMDS at focal gene-regulatory regions, such as transcription start sites (TSSs) and transcription factor binding sites in individual patients. Although our results suggest a potentially distinct fragmentation program near telomeric regions during immunotherapy—potentially driven by chromatin organization, genomic instability, or differences in cell death mechanisms between patient groups—the limited sequencing depth precluded higher-resolution exploration of rMDS in these regions. However, our empirical downsampling analysis demonstrates that the biological signal captured by rMDS is robust and not exclusively dependent on high-depth sequencing. While deeper sequencing may enhance precision and reduce variance at critical gene-regulatory elements, the discriminatory structure underlying rMDS remains robust at the coverage levels utilized in this study. Similarly, although Cluster 1 was enriched for keratin-associated loci, the low coverage and lack of complementary gene expression or methylation data in matched tissues prevented definitive attribution of observed changes to epithelial-specific processes.

Our analysis also faced challenges related to pronounced batch effects. To address this, we trained a batch correction model using plasma collection site, cfDNA isolation date, and WGS sequencing date as covariates, while explicitly preserving the component associated with treatment response. This approach was designed to remove technical variation while maintaining the rMDS signal’s discriminatory power. Although the batch correction model was fit on the full analysis set, including all institutes, it is important to note that the pre-holdout set was not included in model fitting. As a result, evaluation in the pre-holdout set remains valid, albeit with lower performance. Importantly, this analysis was included to demonstrate the effect of incomplete batch correction on fragmentomic predictors rather than to serve as the primary measure of model performance. Future work should focus on developing improved batch correction strategies specifically tailored for cfDNA fragmentation data. Additionally, further validation in independent cohorts, ideally with deeper cfDNA WGS coverage, is a critical next step.

Moreover, our reliance on engineered features—such as the arbitrary 4-mer motif length at the 5′ end—while interpretable, may underutilize the rich sequence context present in cfDNA fragments. Recent studies have suggested that transformer-based models, which capture long-range dependencies and positional information in DNA, could be valuable in this context. Future work should explore the use of attention-based architectures trained directly on cfDNA sequences, as these may reveal informative embeddings beyond motif frequency or entropy, especially in low-depth sequencing scenarios.

Finally, although our results were validated across hold-out splits and multiple timepoints, validation in external cohorts with alternative therapeutic regimens will be necessary to fully elucidate patient response mechanisms to immunotherapy. All patients in this study received neoadjuvant pembrolizumab and underwent surgery, which may limit generalizability to recurrent/metastatic cases and other clinical settings. Prospective validation in independent cohorts, particularly those treated with different immunotherapy regimens, will be valuable. We also note that the prognostic utility of rMDS appears primarily suited for short-term response prediction rather than long-term survival. Although rMDS served as a robust indicator of DFS, its association with OS did not reach statistical significance, warranting a more moderated interpretation of its broader prognostic applicability. This distinction may partly reflect the current follow-up duration as well as the potential confounding effects of subsequent lines of therapy on OS. Importantly, early-event endpoints are increasingly recognized as clinically meaningful measures of therapeutic benefit. For example, the recent FDA approval of neoadjuvant and adjuvant pembrolizumab for resectable locally advanced head and neck squamous cell carcinoma (KEYNOTE-689)^20^ was based on improvements in event-free survival (EFS)—a composite endpoint encompassing disease recurrence, progression preventing definitive surgery, or death—rather than OS. In a similar conceptual framework, DFS served as the primary endpoint in our study and represents an analogous early measure of treatment outcome. Consequently, while rMDS shows promise for early risk stratification based on DFS, further validation in larger cohorts with extended follow-up will be necessary to determine its definitive role in predicting long-term survival outcomes.

In conclusion, this study establishes cfDNA rMDS as a biologically meaningful and clinically predictive biomarker for immunotherapy response in HNSCC. By introducing rMDS as a temporally dynamic signal, we provide a framework for stratifying patients based on treatment response, with the potential to inform and personalize clinical decision-making. Future studies are warranted to evaluate rMDS in broader cancer types, to integrate it with other multimodal biomarkers, and to assess its clinical utility in real-time therapeutic settings.

## Methods

### Study cohort and sample collection

This prospective, multi-institutional Phase II clinical trial (NCT02641093) enrolled patients with newly diagnosed, surgically resectable, locally advanced HNSCC receiving neoadjuvant and adjuvant immunotherapy. The study was approved by the institutional review boards of all participating sites and conducted in accordance with the Declaration of Helsinki and Good Clinical Practice guidelines. Written informed consent was obtained from all participants prior to study procedures.

Patients were recruited from six academic centers: University of Cincinnati (UC), University of Michigan (U-M), Ohio State University (OSU), Medical University of South Carolina (MUSC), MD Anderson Cancer Center (MDACC), and University of Louisville (UofL). Each patient received a single intravenous dose of pembrolizumab (200 mg) 7–21 days prior to surgery, followed by adjuvant radiotherapy (60–66 Gy in 30–33 fractions). High-risk patients received concurrent weekly cisplatin (40 mg/m²), and all patients were eligible for adjuvant pembrolizumab (200 mg IV every 3 weeks, up to six doses).

Eligibility criteria included age ≥18 years, newly diagnosed and histologically or cytologically confirmed HNSCC, and locally advanced disease classified as stage III or IV according to the AJCC 8th edition (T3 or T4 tumor, ≥N2 nodal disease, or clinical evidence of extranodal extension (ENE) on imaging). Tumors had to be deemed resectable by the treating head and neck surgeon, with no involvement of the skull base or T4b stage. Additional inclusion criteria were an Eastern Cooperative Oncology Group (ECOG) performance status ≤1 and adequate organ function.

Key exclusion criteria were human papillomavirus-positive (HPV+) oropharyngeal cancer (HPV+ disease outside the oropharynx was permissible, although HPV testing was not mandatory), nasopharyngeal cancer, metastatic disease (as determined by chest CT or PET/CT), autoimmune disease, active intercurrent illness (including significant cardiovascular disease, active viral infections, or major psychiatric illness), and steroid use exceeding prednisone 10 mg daily. For comprehensive details, refer to the original study^19^.

Peripheral blood collected at the first three timepoints was used in this study: (i) pre-treatment screening (Screen), (ii) immediately after surgical resection (Day 0), and (iii) approximately 3–10 weeks following surgery (Adjuvant Week 1). We included all plasma samples available from the trial for cfDNA analysis. Samples for which only whole blood was collected were excluded, as genomic DNA contamination in whole-blood specimens can compromise the integrity of cfDNA fragmentation measurements.

### Treatment response and pathological risk determination

Pathological response to neoadjuvant pembrolizumab was evaluated by centralized histopathological analysis. Resected tumor specimens were assessed by experienced pathologists for standard pathological risk features, including extracapsular nodal extension (ENE), margin status, lymphovascular invasion (LVI), perineural invasion (PNI), and the number of involved lymph nodes. For comprehensive details, refer to the original study^19^.

The primary metric for treatment response was pathological treatment effect (TE), defined as tumor necrosis accompanied by histiocytic inflammation and/or giant cell reaction to keratinaceous debris. TE was quantified as the ratio of the area showing these features to the total area comprising residual viable tumor and TE. Pathological response was classified as no response (TE <10%) or response (TE ≥10%), similar to previous studies^20,41^. The presence of positive surgical margins and/or extranodal extension (ENE) was used to define pathological risk. For comprehensive details, refer to the original study^19^.

### Sex as a biological variable

Our study examined male and female humans, and there is no significant impact of sex on the conclusion.

### Plasma processing and cfDNA extraction

Peripheral blood was shipped overnight, and cfDNA was isolated from 500 μL of plasma, with all samples stored at –80°C prior to processing. Before isolation, plasma samples were thawed on ice, centrifuged at 1,600 x g for 10 minutes at 4°C, and then subjected to a second centrifugation at 16,000 x g for 10 minutes at 4°C to remove residual cell debris. cfDNA extraction was performed using the MagMAX Cell-Free DNA Isolation Kit (Applied Biosystems) following the manufacturer’s protocol. cfDNA concentration and size distribution were measured using Qubit (Invitrogen) and BioAnalyzer (Agilent), respectively. For each patient, plasma samples collected at three different time points were processed together within the same isolation batch.

### cfDNA WGS

cfDNA samples were randomly allocated into eight library preparation batches, ensuring that all time points from each patient were included in the same batch. Library preparation was performed on 1[ng of cfDNA using the KAPA HyperPrep Kit (Roche) and NEXTFLEX Unique Dual Index Barcodes (PerkinElmer; 300 nM final concentration). Sequencing was carried out on the Illumina NovaSeq 6000 S4 platform using paired-end 150 bp reads (PE150).

### Data preprocessing of cfDNA WGS data

Raw sequencing data were processed using the FinaleDB Workflow^42^ (https://github.com/epifluidlab/finaledb_workflow), implemented via Snakemake (v7.8.0)^43^. Specifically, paired-end FASTQ files underwent adapter trimming with Trimmomatic (v0.39)^44^, followed by alignment to the human reference genome (hg38) using BWA-MEM (v0.7.15)^45^ with default parameters. PCR duplicates were removed with samblaster (v0.1.24)^46^. Reads were retained if both ends were uniquely mapped, properly paired, non-supplementary, and had mapping quality ≥30. Only autosomal fragments 50-350 bp in length were included. Reads overlapping ENCODE blacklist regions^47^ (hg38) were excluded to eliminate mapping artifacts.

### Regional motif diversity score (rMDS)

Autosome genome was divided into 500kb non-overlapped bins. For each 500 kb bin *j*, the raw rMDS was defined as the normalized Shannon entropy of 5′ end 4-mer motifs:

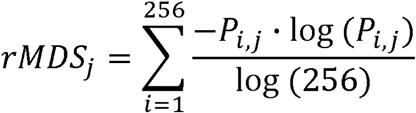

where *P_i,j_* is the relative frequency of the *i_th_* 4-mer end motif in bin *j*. Normalization by log(256) ensures that the score ranges from 0 (minimum diversity) to 1 (maximum diversity). The resulting rMDS values were then z-score transformed across all bins in each sample:

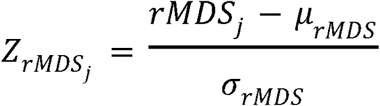

where *µ_TMDS_* and *σ_TMDS_* denote the mean and standard deviation of rMDS values across the genome, respectively. These z-transformed values were used in all downstream analyses. The 500 kb bin size provided a balance between genomic resolution and fragment count per bin (∼2 × 10[fragments/bin at 1.9× coverage).

### Other cfDNA fragmentation feature extraction

Additional fragmentation features were extracted from pre-processed BED and BAM files using FinaleToolkit (v0.10.7)^21^. The autosomal genome was divided into non-overlapping 500 kb bins using Bedtools (v2.31.1)^48^, excluding low-mappability regions identified by the UCSC 45-mer mappability track (hg38)^49^. Fragment-level BED files were used to calculate: (1) fragment length distributions, (2) 4-mer fragment end motif frequencies, (3) global motif diversity scores (MDS), and (4) DELFI scores (with the --no-merge-bins option). GC-corrected coverage values were derived from DELFI-corrected fragment counts. For copy number and tumor fraction analyses, BAM files were processed using ichorCNA (v0.2.0)^22^ with hg38 genome settings at the same resolution.

### Batch correction and dimensionality reduction

Batch correction of rMDS values and other fragmentomic features was performed using a linear model adapted from the limma framework^50^. For each 500 kb genomic bin, rMDS values were modeled as a function of cfDNA isolation date, WGS library preparation date, and collection institute, with treatment response (Responder vs. Non-Responder) included as a preserved biological covariate. Design matrices employed sum contrasts via the *patsy.contrasts.Sum* method (patsy v0.5.6). Linear coefficients were estimated on the analysis set (including the Institute Hold-Out set) using ordinary least squares, and batch effects were regressed out. To preserve the independence of the pre-holdout set, batch correction was performed by applying the design matrix contrasts and regression coefficients learned from the analysis set. No parameters were re-estimated using pre-holdout data. Matrix operations were implemented with NumPy (v1.26.4)^51^ and pandas (v2.2.2)^52^.

Corrected matrices were standardized using z-score normalization (StandardScaler, scikit-learn v1.5.2)^53^. Dimensionality reduction was conducted using truncated singular value decomposition (SVD; *numpy.linalg.svd*)^51^, with components selected based on the singular value spectrum and variance explained (**Supplementary Fig. 5**).

Low-dimensional embeddings were generated using UMAP (umap-learn v0.5.7; metric=’cosine’, n_neighbors=50, min_dist=0.5)^54^.Confidence ellipses (2 SD) for each group were constructed from the eigen decomposition of the covariance matrix, and ellipse intersections were computed using Shapely (v2.1.0). Visualizations were produced with Matplotlib (v3.8.3)^55^ and Seaborn (v0.13.2)^56^. To quantify the quality and stability of the resulting clusters, specifically the separation between responders and non-responders, we calculated Silhouette scores. The Silhouette score for each sample was computed as *(b−a)/max(a,b),* where *a* is the mean intra-cluster distance and *b* is the mean distance to the nearest neighboring cluster.

### Sequencing Depth and Stability Analysis

To evaluate the impact of sequencing depth on rMDS, we used a reference cfDNA WGS sample from BH01 in Snyder et al. 2016^13^ (56x effective coverage) and performed empirical downsampling to 50x, 25x, 10x, 5x, 2x, 1x, and 0.1x effective coverage. Pearson correlation coefficients were computed between each downsampled profile and the 56x reference to assess signal recovery. Additionally, responder and non-responder samples were downsampled to assess the stability of group-level biological discrimination across varying depths.

### Differential analysis of rMDS

Differential analysis of rMDS between treatment responders and non-responders was performed using the limma^50^ package in R with a linear mixed-effects modeling approach to account for repeated measures across timepoints. A global design matrix incorporating treatment response group and clinical covariates (age, race, gender, ethnicity, diagnosis, smoking, alcohol use) was constructed, with within-patient correlation estimated using patient ID as a blocking variable and array-specific weights applied to handle variance heterogeneity. Linear models were fitted using empirical Bayes moderation, and raw p-values were corrected for multiple testing using q-values to control the false discovery rate. Genomic bins with q-values < 0.1 were considered significantly differential between responders and non-responders.

### Clustering and chromosomal enrichment of differential rMDS regions

Using clinical covariate-corrected (Supplemental Figure 9) and uncorrected rMDS data (Figure 3), differential 500 kb bins were clustered based on rMDS values across the three timepoints (Screen, Day 0, Adjuvant Week 1). Genomic bins (columns) were hierarchically clustered using Ward’s method (*scipy.cluster.hierarchy.linkage*)^57^, with clusters determined via fcluster (t=3, criterion=’maxclust’). Patient rows were independently clustered within each treatment group. Heatmaps were generated with Marsilea (v0.5.1)^58^.

For each cluster, the average rMDS trajectory across timepoints was computed and stratified by response group. Longitudinal patterns were visualized as line plots (Seaborn v0.13.2, ci=95)^56^.

Telomeric enrichment was assessed by calculating the distance from each bin midpoint to the nearest chromosome end using hg38.chrom.sizes (UCSC)^49^. For each cluster, the empirical distance distribution was compared to a null distribution generated from 100,000 randomly sampled 500 kb bins, using a one-sided Mann–Whitney U test (*scipy.stats.mannwhitneyu*)^57^ and a permutation test to determine whether observed distances were significantly shorter than expected. Density plots were generated with kernel density estimation (*scipy.stats.gaussian_kde*)^57^, normalized for comparison.

### Gene ontology and pathway enrichment analysis of differential rMDS regions

Functional enrichment analysis was performed for each rMDS cluster using g:Profiler (version e112_eg59_p19_25aa4782)^59^. Genomic bins were mapped to nearby genes using default Ensembl annotations. Enrichment of GO Biological Process and Reactome pathway terms was tested, and fold enrichment was calculated as (intersection size / query size) divided by (term size / effective domain size). The top 10 significant enriched terms with highest fold changes were visualized in the main figure.

### Locus-specific visualization of differential rMDS signal profiles

Genome-wide sliding windows of 500 kb (step size: 10 kb) were generated. Basically, fragment end motif profiles were computed using the interval-end-motifs and rmds modules from FinaleToolkit (v0.10.7)^21^ in each 10kb window by using the rMDS value within the nearby 500kb window (± 250kb from the center of 10kb window). Signal matrices were z-score normalized, batch-corrected, and averaged by group (response and timepoint). Visualization was performed using IGV (v2.19.2)^60^, employing color-coded overlays to differentiate response groups and timepoints.

### Machine learning classification of treatment response

To evaluate the predictive power of rMDS features, models were trained using TabPFNClassifier (tabpfn v2.0.8)^37^. Input matrices were batch-corrected, z-score normalized, and reduced via truncated SVD (scikit-learn v1.5.2, n_components=6)^53^. Model performance was assessed using ROC AUC, accuracy, F1 score, precision, recall, and balanced accuracy. Patient-level predictions were obtained by using the most recent available time point (Adjuvant Week 1 or Day 0).

Model robustness was tested to evaluate the effect of random sampling by three approaches:

- Cross-Validation Evaluation: The model underwent 100 rounds of 10-fold cross-validation at the patient level. In each round, patients were randomly divided into 10 groups, with 9 groups used for training and 1 group held out for testing. This process was repeated 10 times per round (rotating which group was held out) and then replicated 100 times with different random splits.
- Single-Institute Holdout Evaluation: The model was trained using data from 5 institutes, then tested separately on data from the held-out institute.
- Pre-Holdout Evaluation: The model underwent 100 rounds of testing, where 10 patients were randomly selected as the test set in each round. Importantly, any batch correction factors were learned only from the training set, ensuring the test remained truly independent.

Performance metrics were computed at both the sample and patient levels. ROC curves and confusion matrices were visualized using Seaborn (v0.13.2)^56^ and Matplotlib (v3.8.3)^55^.

### Survival analysis

Survival analyses were performed in R (v4.5.0) using survival (v3.8.3)^61,62^. Kaplan–Meier estimates for disease-free survival (DFS) and overall survival (OS) were calculated, with time-to-event defined as days from the Adjuvant Week 1 dose. Patients missing this timepoint were excluded from downstream analysis.

PD-L1 expression was assessed using immunohistochemistry (IHC). Formalin-fixed, paraffin-embedded tumor tissue samples were stained using the 22C3 antibody clone (Agilent, Dako) and evaluated with the FDA-approved PD-L1 IHC pharmDx assay. Testing was performed at Caris Life Sciences or NeoGenomics Laboratories and independently confirmed by a board-certified pathologist at the University of Cincinnati. A combined positive score (CPS) was calculated as: CPS = (Number of PD-L1–positive tumor cells + PD-L1–positive immune cells)*100 / (Total number of viable tumor cells). For comprehensive details, refer to the original study^19^.

Survival analyses were stratified by predicted treatment response (Responder vs. Non-Responder) from the single-institute hold-out evaluation, pathological risk categories (Intermediate vs. High), PD-L1 CPS categories (>20, 1-19, 0), and tumor fraction (Low vs. High, dichotomized at the median). Kaplan-Meier survival curves were generated with accompanying risk tables showing the number of patients at risk. Log-rank tests were applied for group comparisons^61^. Visualizations were created using ggplot2 (v3.5.2)^63^, with pairwise comparisons performed for multi-level groups.

Cox proportional hazards regression models were fitted to estimate hazard ratios (HR) with 95% confidence intervals. Univariate analyses were performed for age, gender, HPV status (determined by p16 immunohistochemistry as a surrogate), smoking history, alcohol use, predicted response, risk stratification, PD-L1 expression, and tumor fraction. Results were considered statistically significant at p < 0.05.

## Data and code availability

All analysis code is available at the study’s GitHub repository: https://github.com/epifluidlab/headneck. Raw whole-genome sequencing data from plasma cfDNA will be available in the European Genome-phenome Archive (EGA) with controlled access under accession code (EGAS50000001286). Data access can be obtained through a request to the corresponding authors. The corresponding authors will generally respond to requests within one week. Once granted, the access has no time restriction The raw sequencing data are protected by data privacy laws. The de-identified post-processed fragment files are available in zenodo.org (doi: 10.5281/zenodo.15738112).

## Acknowledgments

Y.L. is supported by the startup grant to Y.L. from Cincinnati Children’s Hospital Medical Center, Northwestern University, Robert H. Lurie Comprehensive Cancer Center of Northwestern University, and NHGRI (R56HG012360 to Y.L.). This work was also supported by a Science Olympiad Alumni Research (SOAR) Grant from the Science Olympiad USA Foundation. Dr. Wise-Draper was funded by Merck Sharp & Dohme Corp., a subsidiary of Merck & Co., Inc., Kenilworth, NJ, USA for the conduct of the clinical trial. Additional support was provided through the computational resources and staff contributions provided for the Quest high performance computing facility at Northwestern University, which is jointly supported by the Office of the Provost, the Office for Research, and Northwestern University Information Technology. This research was also supported in part through the computational resources and staff contributions provided by the Genomics Compute Cluster, which is jointly supported by the Feinberg School of Medicine, the Center for Genetic Medicine, and Feinberg’s Department of Biochemistry and Molecular Genetics, the Office of the Provost, the Office for Research, and Northwestern Information Technology. The Genomics Compute Cluster is part of Quest, Northwestern University’s high performance computing facility, with the purpose to advance research in genomics.

## Author Contributions

Y.L. and T.W.-D. conceived the study. H.F. and L.W. performed the cfDNA extraction and library constructions. R.B and H.Z. performed the data analysis with input from Y.L, T.W.-D., S.G., M.T., B.Z. M.K., D.L., and D.A.H.. F.P.W, M.O.O, N.E.D, J.M.K, M.G., D.A.E., and T.W.-D. Designed the clinical trial and provided the blood samples. R.B. and Y.L wrote the manuscript together. All authors read and approved the final manuscript.

