## Supplementary Materials for "Genome-Wide Variations of End Motif in Cell-Free DNA Fragments Distinguish Immunotherapy Responders from Non-Responders in Head and Neck Cancer: A Multi-Institute Prospective Study"

### Supplemen0tary Figures

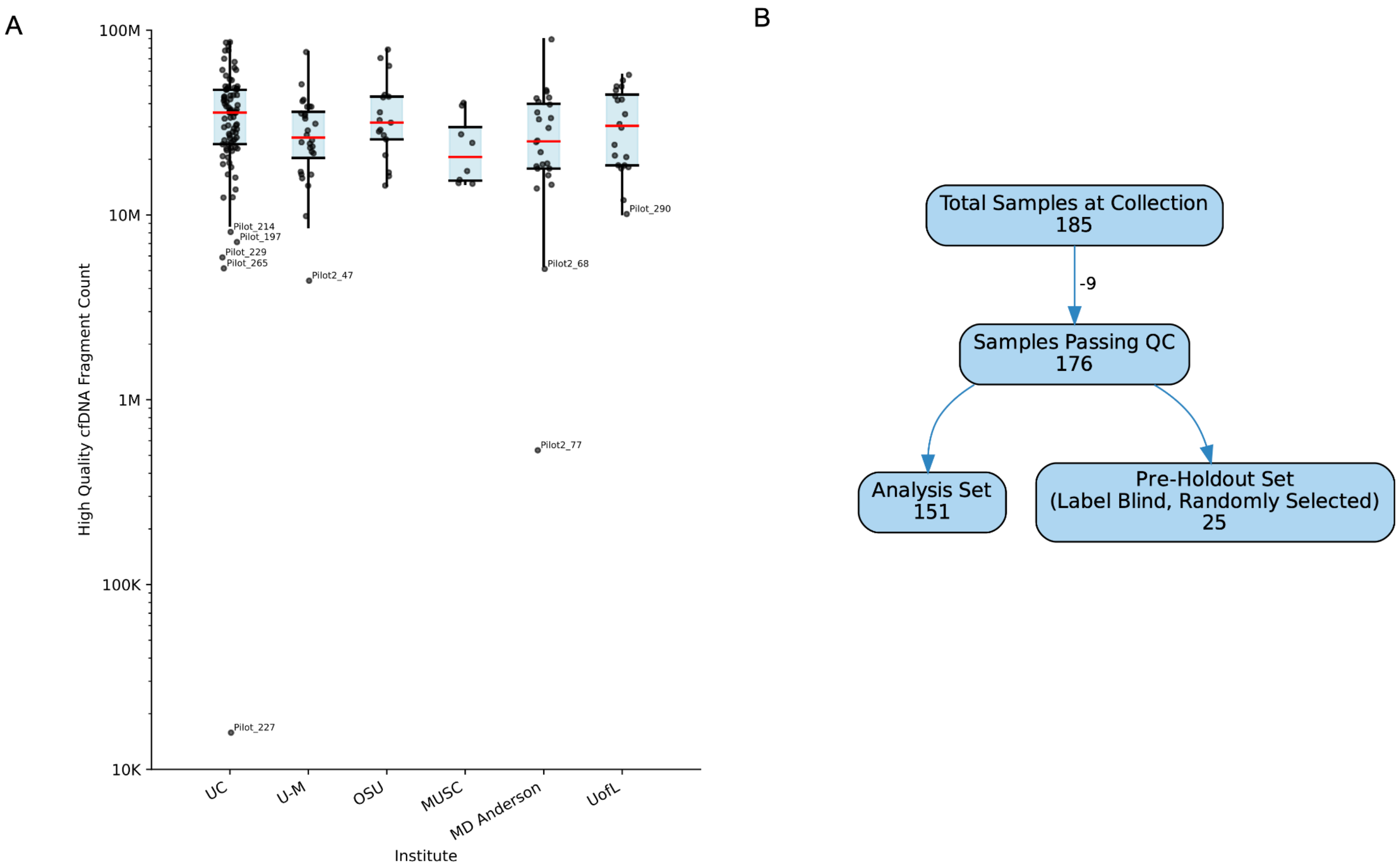


**Supplementary Figure 1. Sample selection workflow and number of high-quality cfDNA fragments across institutes. (A)** Distribution of high-quality cfDNA fragment counts (MAPQ>30, no ENCODE Blacklist overlap) per sample ($\log_{10}$ scale) stratified by collection site: University of Cincinnati (UC), University of Michigan (U-M), Ohio State University (OSU), Medical University of South Carolina (MUSC), The University of Texas MD Anderson Cancer Center and University of Louisville (UofL). Boxplots show the interquartile range with whiskers at 1.5*Interquartile Range, red lines indicate medians, and individual dots represent samples (extreme outliers are labeled). **(B)** Flowchart of plasma sample processing: of 185 initially collected samples, 9 failed quality control due to low number of reads. From these QC-passed samples, 151 were allocated to the primary analysis cohort, while 25 (10 patients) were randomly set aside as a label-blinded pre-holdout subset.

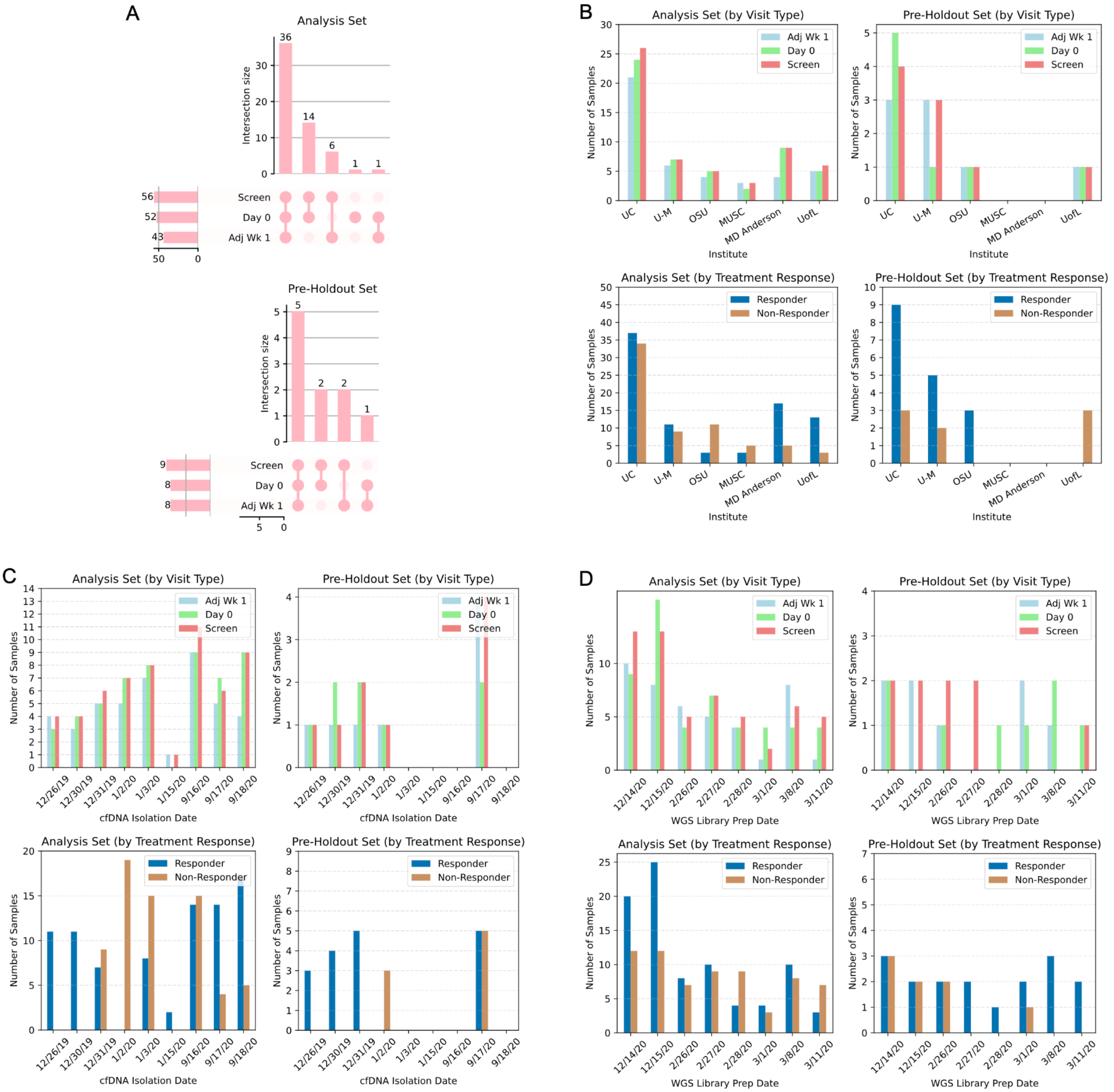


**Supplementary Figure 2. Distribution of available plasma samples across clinical time points, institutions, cfDNA isolation dates, and WGS library preparation dates. (A)** UpSet plots showing the overlap of available samples at Screening, Day 0, and Adjuvant Week 1 visits for the analysis and pre-holdout sets. **(B)** Number of plasma samples from each participating institution, separated by visit type, treatment response and set. **(C)** Distribution of cfDNA isolation dates across visit types and treatment response for the analysis and pre-holdout sets. **(D)** Distribution of WGS library preparation dates across visit types and treatment response for the analysis and pre-holdout sets.

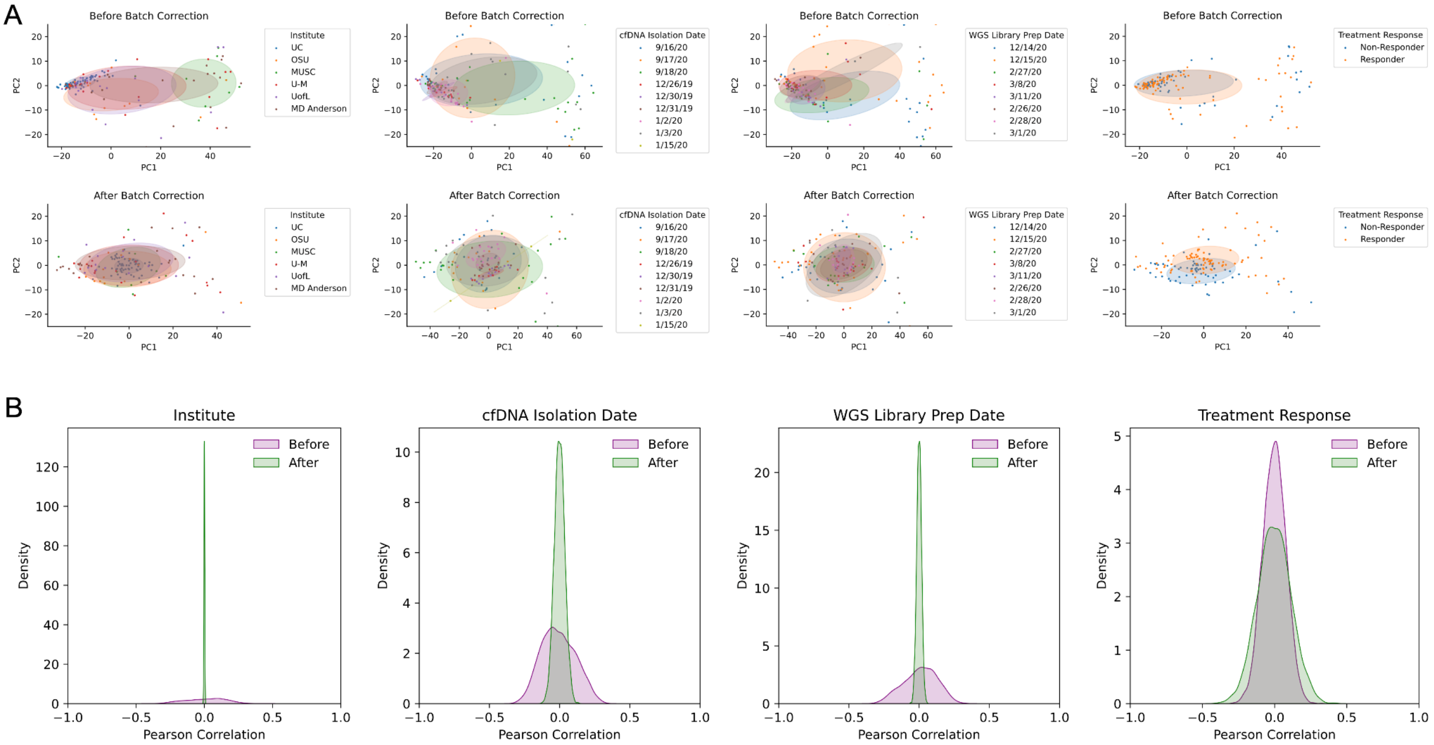


**Supplementary Figure 3. Evaluation of batch effect correction on rMDS values across all plasma samples. (A)** PCA plots of rMDS values colored by metadata variables before (top row) and after (bottom row) batch correction. Ellipses denote ~1 standard deviation contours along the centroid of the first two principal components. Correction reduces visible clustering by batch variables while maintaining separation by treatment response. **(B)** Density plots of Pearson correlation coefficients between rMDS value in each 500kb genomic bin and possible batch effect variables or labels--plasma collection institute, cfDNA isolation date, WGS library preparation date, and treatment response--before (purple) and after (green) batch correction. Batch correction successfully eliminates correlations with technical factors while preserving biological signals related to treatment response.

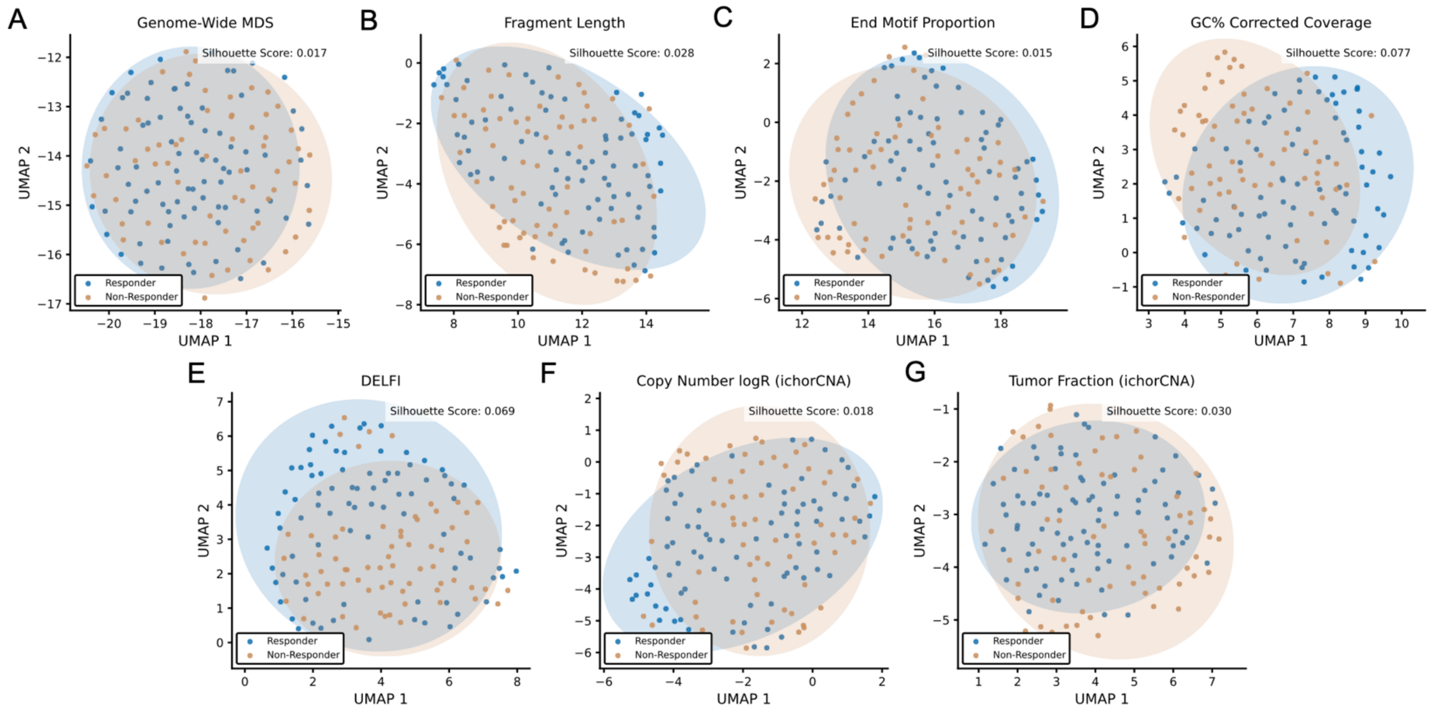


**Supplementary Figure 4. UMAP plot of cfDNA fragmentation-derived features by treatment response.** Uniform Manifold Approximation and Projection (UMAP) plots of cfDNA-derived fragmentation features, colored by treatment response status (blue: Responder, brown: Non-Responder). Each panel represents UMAP clustering based on a distinct feature: **(A)** genome-wide MDS, **(B)** fragment length, **(C)** 5' end motif proportion, **(D)** GC%-corrected fragment coverage, **(E)** DELFI score, **(F)** copy number logR (ichorCNA), **(G)** tumor fraction (ichorCNA). Ellipses indicate ~2 standard deviations from the centroid of each group.

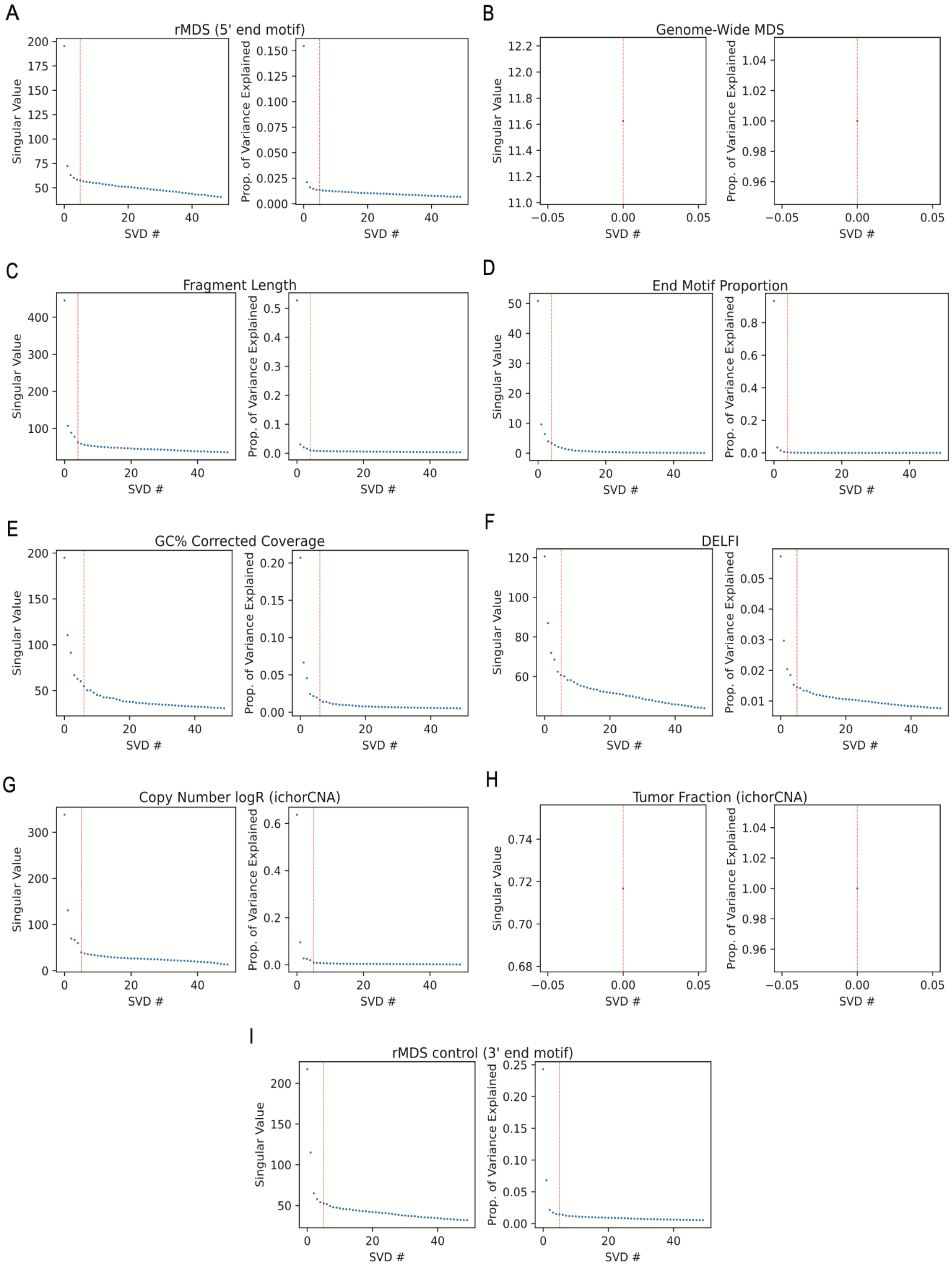


**Supplementary Figure 5. Selection of top singular value decomposition (SVD) components for UMAP.** Singular value decomposition (SVD) spectra for each cfDNA-derived feature set used in UMAP analysis. Left panels show the singular value spectrum, and right panels display the cumulative variance explained. Vertical red lines indicate the selected number of SVD components used for UMAP embedding. Panels: **(A)** rMDS (5’end), **(B)** genome-wide MDS, **(C)** fragment length, **(D)** end motif proportion, **(E)** GC%-corrected coverage, **(F)** DELFI score, **(G)** copy number logR (ichorCNA), **(H)** tumor fraction (ichorCNA), and **(I)** rMDS control (from 3’ end 4-mer motif).

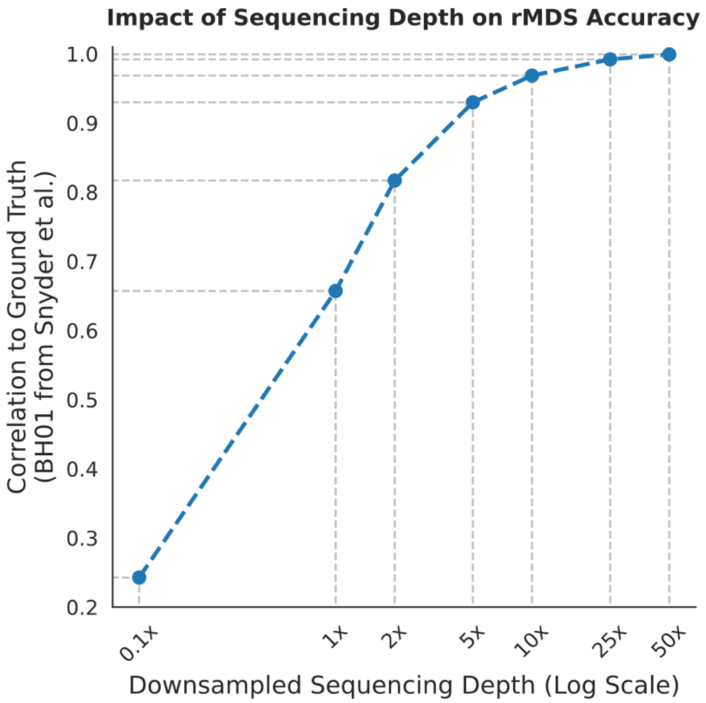


**Supplementary Figure 6. rMDS estimation stability across varying sequencing depths.** Evaluation of rMDS profile stability using the deeply sequenced non-pregnant healthy (BH01) cfDNA WGS sample from Snyder et al. (56x effective coverage, ~96x raw coverage) downsampled to various depths (0.1x to 50x).

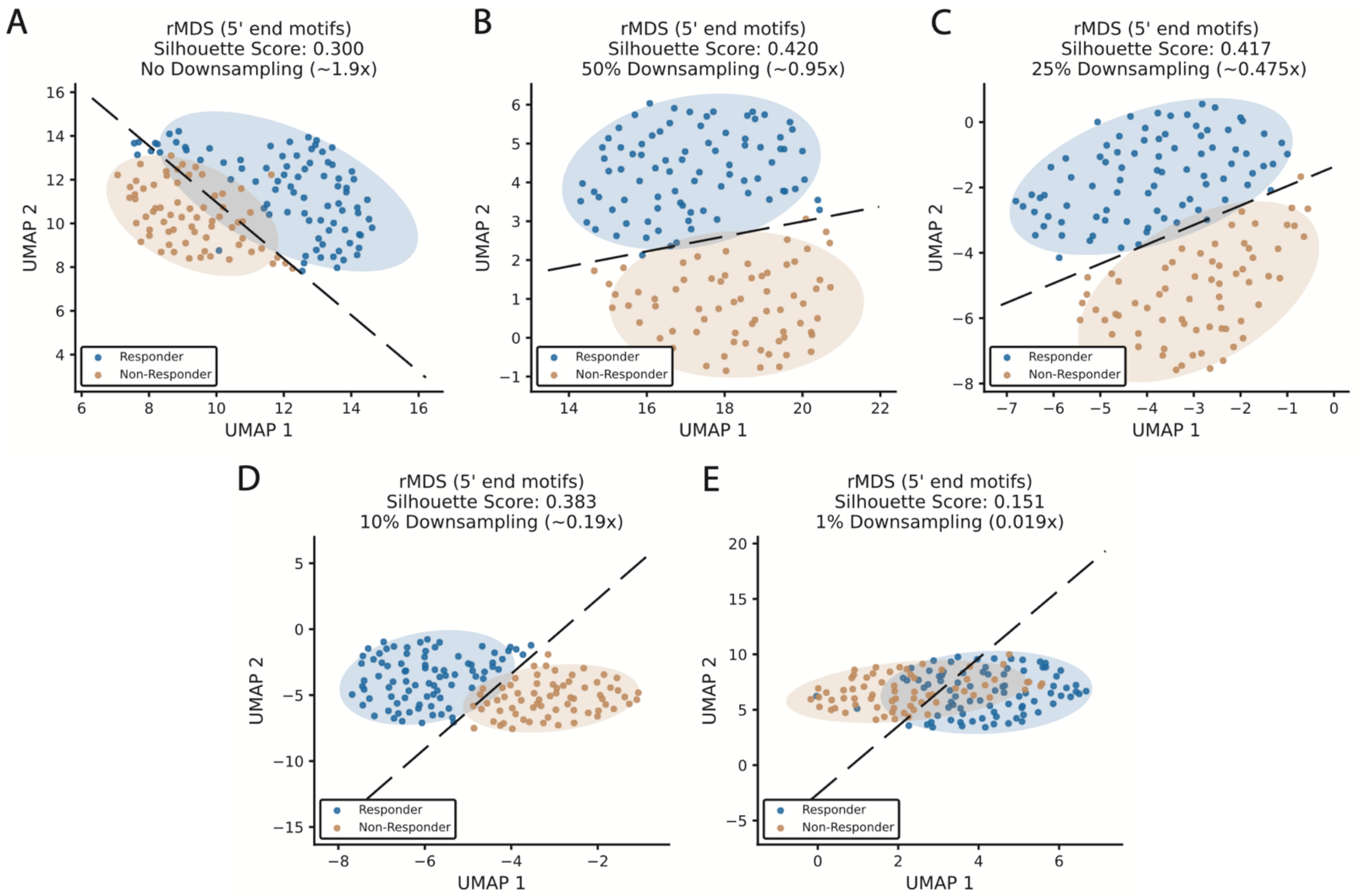


**Supplementary Figure 7. Group-level discrimination of responders and non-responders across downsampled sequencing depths.** UMAP projections of regional motif deviation scores (rMDS) from 5' end motifs at various downsampling levels (ranging from **(A)** no downsampling [~1.9x], **(B)** 50% downsampling [~0.95x], **(C)** 25% downsampling [~0.475x], **(D)** 10% downsampling (~0.19x), to **(E)** 1% downsampling [~0.019x]). Blue and orange ellipses represent Responders and Non-Responders, respectively.

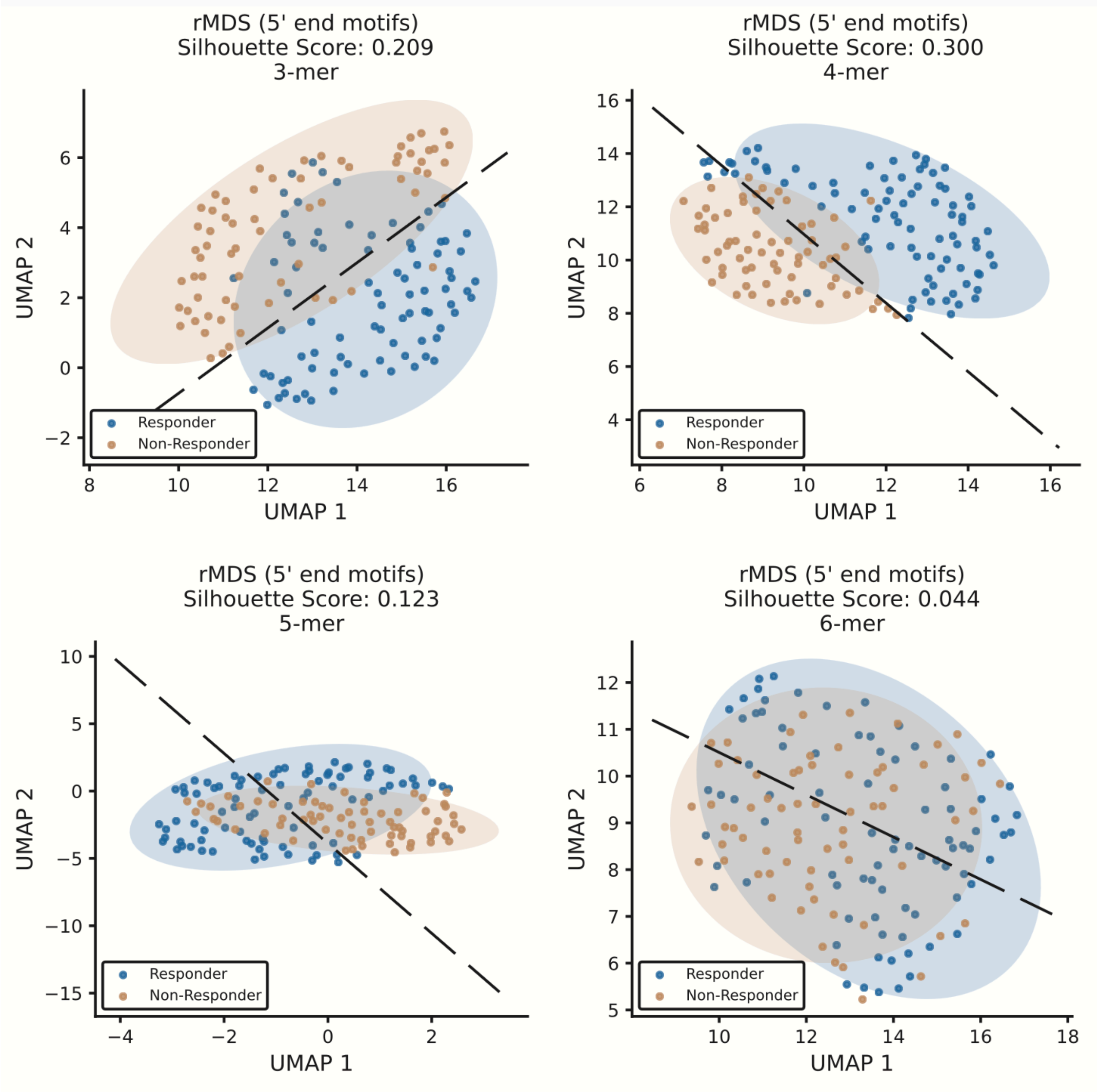


**Supplementary Figure 8. Comparison of group-level discrimination across different k-mer lengths for rMDS.** UMAP projections of regional motif deviation scores (rMDS) using **(A)** 3-mer, **(B)** 4-mer, **(C)** 5-mer, and **(D)** 6-mer 5' end motifs.

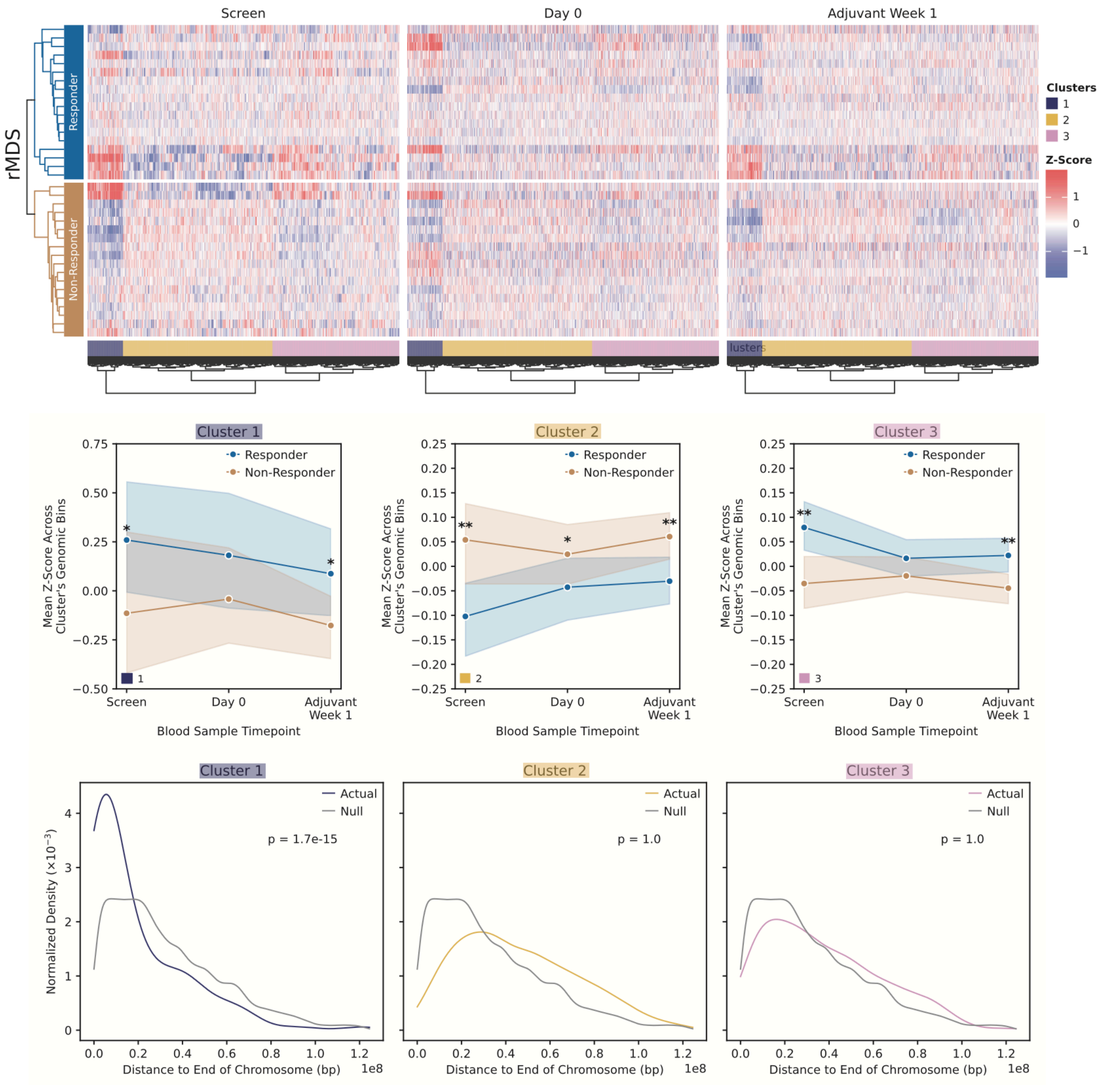


**Supplementary Figure 9. Differential rMDS regions between immunotherapy responders and non-responders using covariate corrected rMDS.** Top panel shows heatmap of differentially enriched rMDS regions across three timepoints (Screen, Day 0, Adjuvant Week 1) reveals distinct longitudinal dynamics between responders and non-responders, stratified by cluster. Middle panel, line plots show the mean z-score trajectory over time for each cluster, separated by response status; p-values indicate statistical significance from one-sided Welch's t-tests comparing Responders and Non-Responders at each timepoint (**p* < 0.1; ***p* < 0.01). Bottom panel shows the normalized density of distances from each rMDS region to the nearest chromosome end for each cluster, compared to a null distribution; p-values indicate one-sided Mann-Whitney U tests assessing whether the actual distribution is significantly closer to chromosome ends than the null.

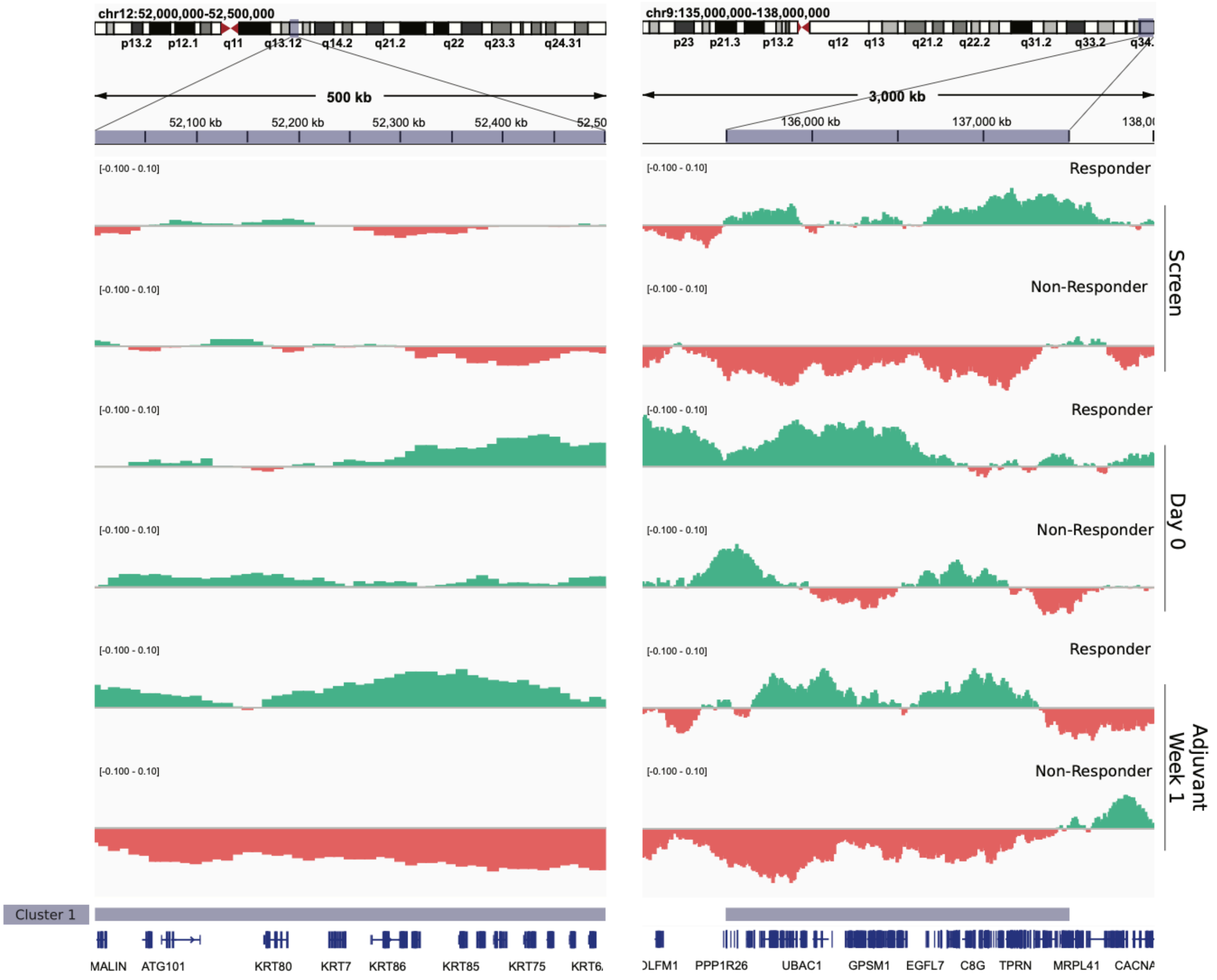


**Supplementary Figure 10. Example of rMDS signal profiles at loci enriched for keratinization genes and telomere-proximal domains.** Genomic regions chr12:52,000,000–52,500,000 (left) and chr9:135,500,000–137,500,000 (right) were selected based on high density of keratinization-associated genes and a large telomere-proximal domain, respectively. Normalized rMDS Z-scores are shown for responders and non-responders at three timepoints (Screen, Day 0, and Adjuvant Week 1). Red and green shading indicate relative depletion and enrichment, respectively, compared to baseline. Relevant gene annotations are provided below the tracks.

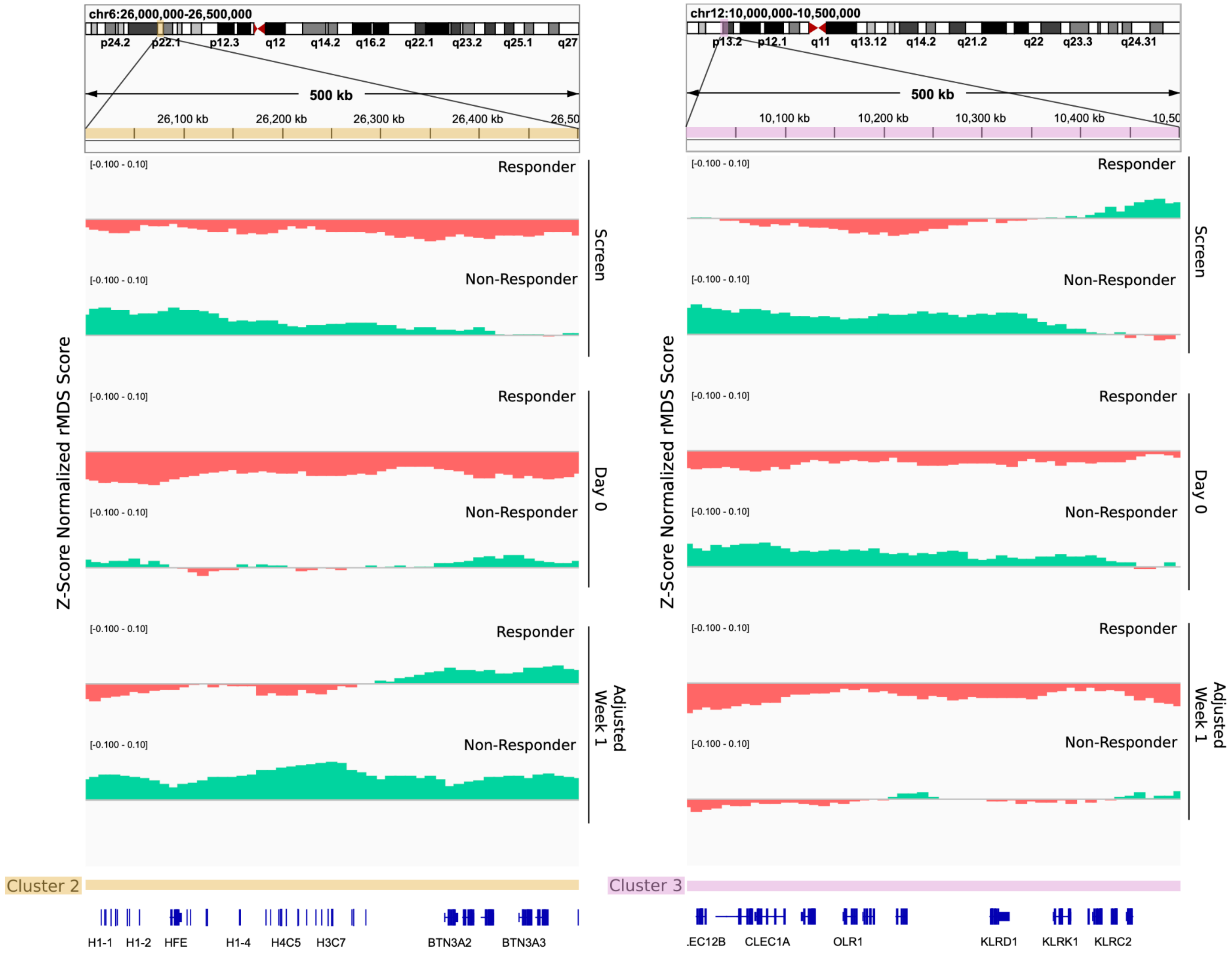


**Supplementary Figure 11. Example rMDS signal profiles at loci enriched for CENP-A and lectin receptor genes.** Genomic regions chr6:26,000,000-26,500,000 (left) and chr12:10,000,000-10,500,000 (right) were selected based on high density of CENP-A related (Cluster 2) and killer lectin-like receptor (Cluster 3) genes, respectively. Normalized rMDS Z-scores are shown for responder and non-responder patients at three timepoints (Screen, Day 0, and Adjuvant Week 1). Red and green shading indicate relative depletion and enrichment, respectively, compared to baseline. Relevant gene annotations are provided below the tracks.

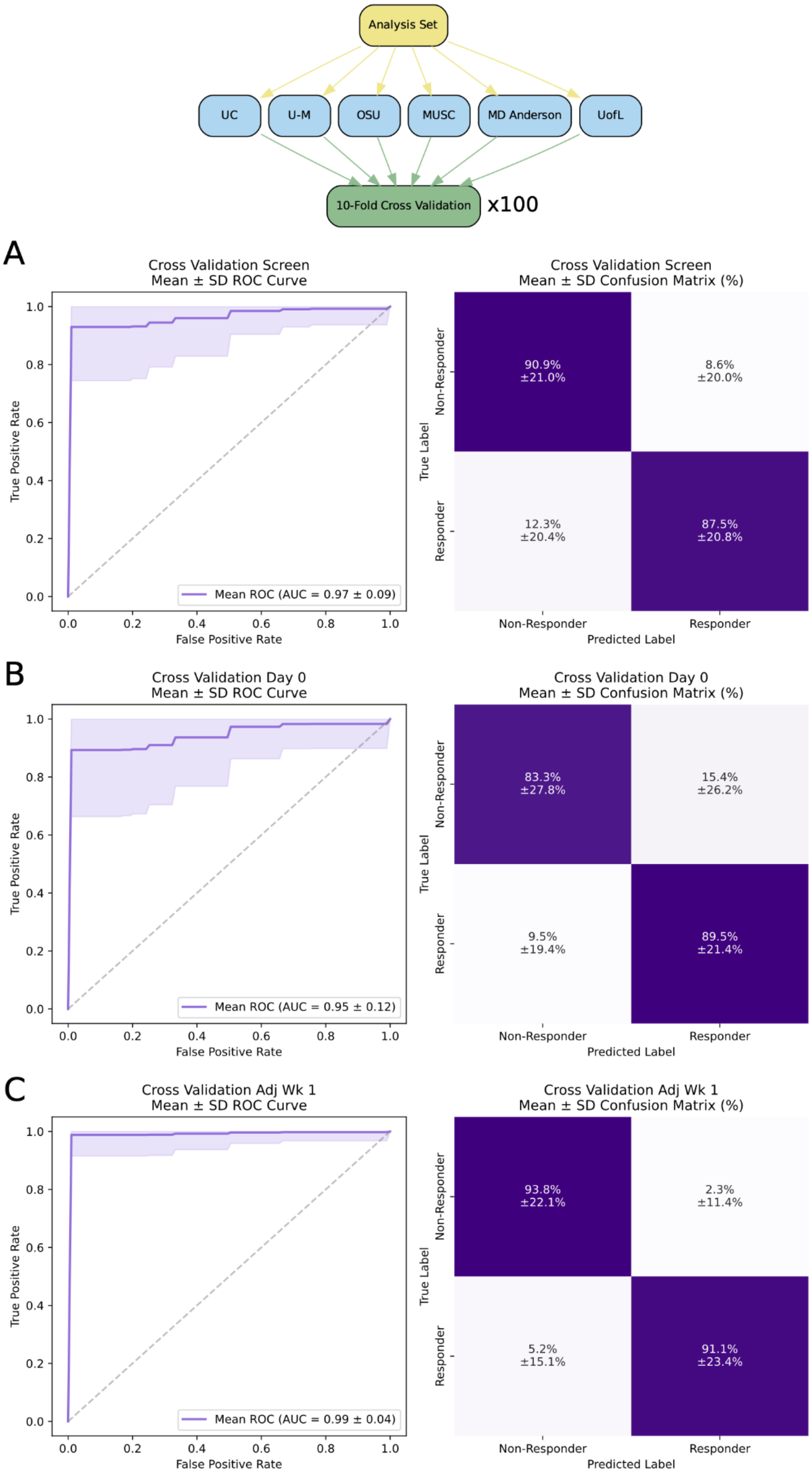


**Supplementary Figure 12. Sample-level performance of rMDS model across clinical timepoints in the cross-validation evaluation strategy.** Receiver operating characteristic (ROC) curves and confusion matrices for the rMDS model evaluated on 10-fold patient-level cross validation repeated 100 times. Performance is shown overall and stratified by plasma collection timepoints: **(A)** Screen, **(B)** Day 0, and **(C)** Adjuvant Week 1. ROC AUC values and their standard deviation are indicated.

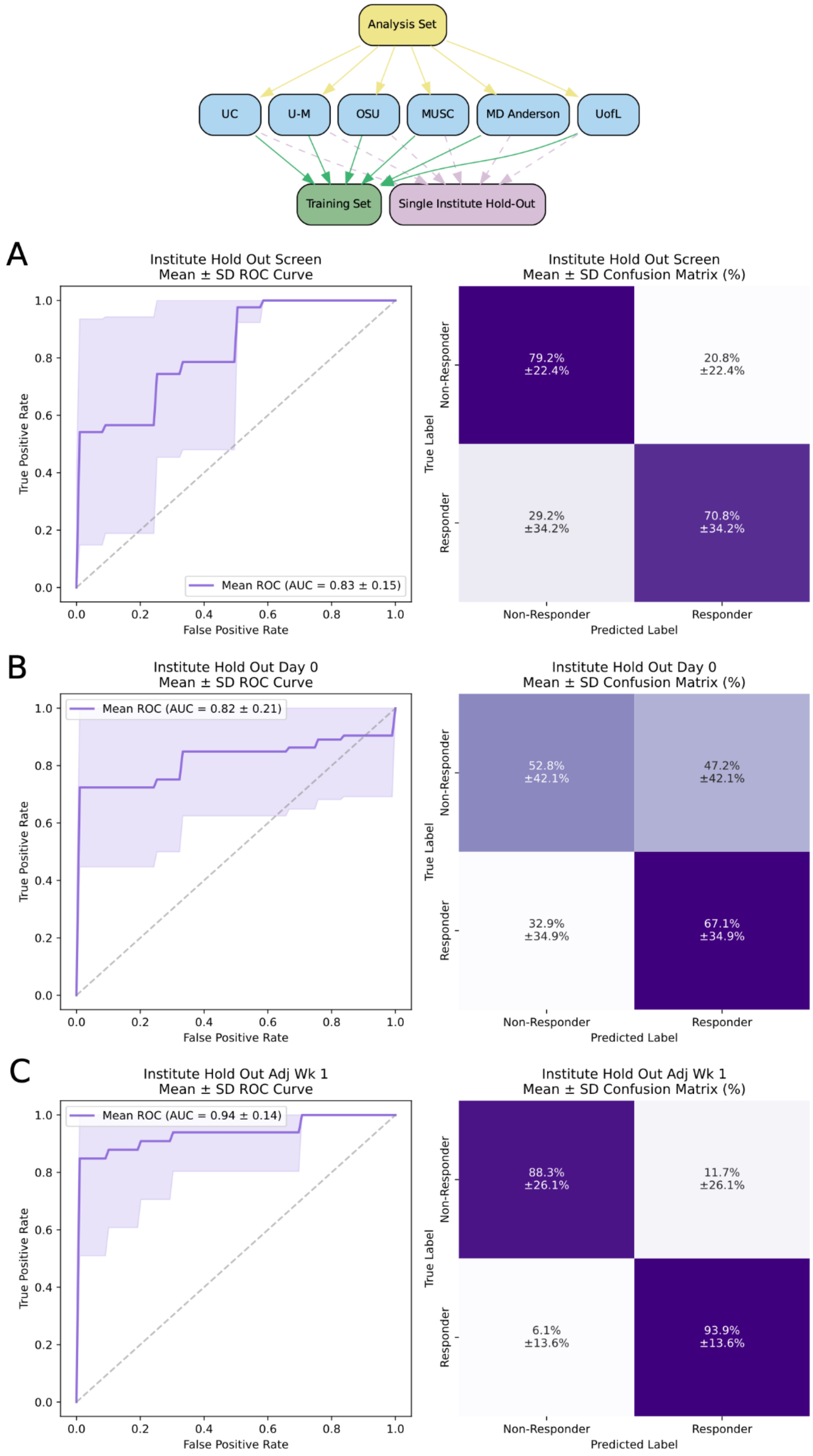


**Supplementary Figure 13. Sample-level performance of rMDS model across clinical timepoints in the institute hold-out evaluation strategy.** Receiver operating characteristic (ROC) curves and confusion matrices for the rMDS model evaluated on single institute hold-out setup. Performance is shown overall and stratified by plasma collection timepoints: **(A)** Screen, **(B)** Day 0, and **(C)** Adjuvant Week 1. ROC AUC values and their standard deviation are indicated.

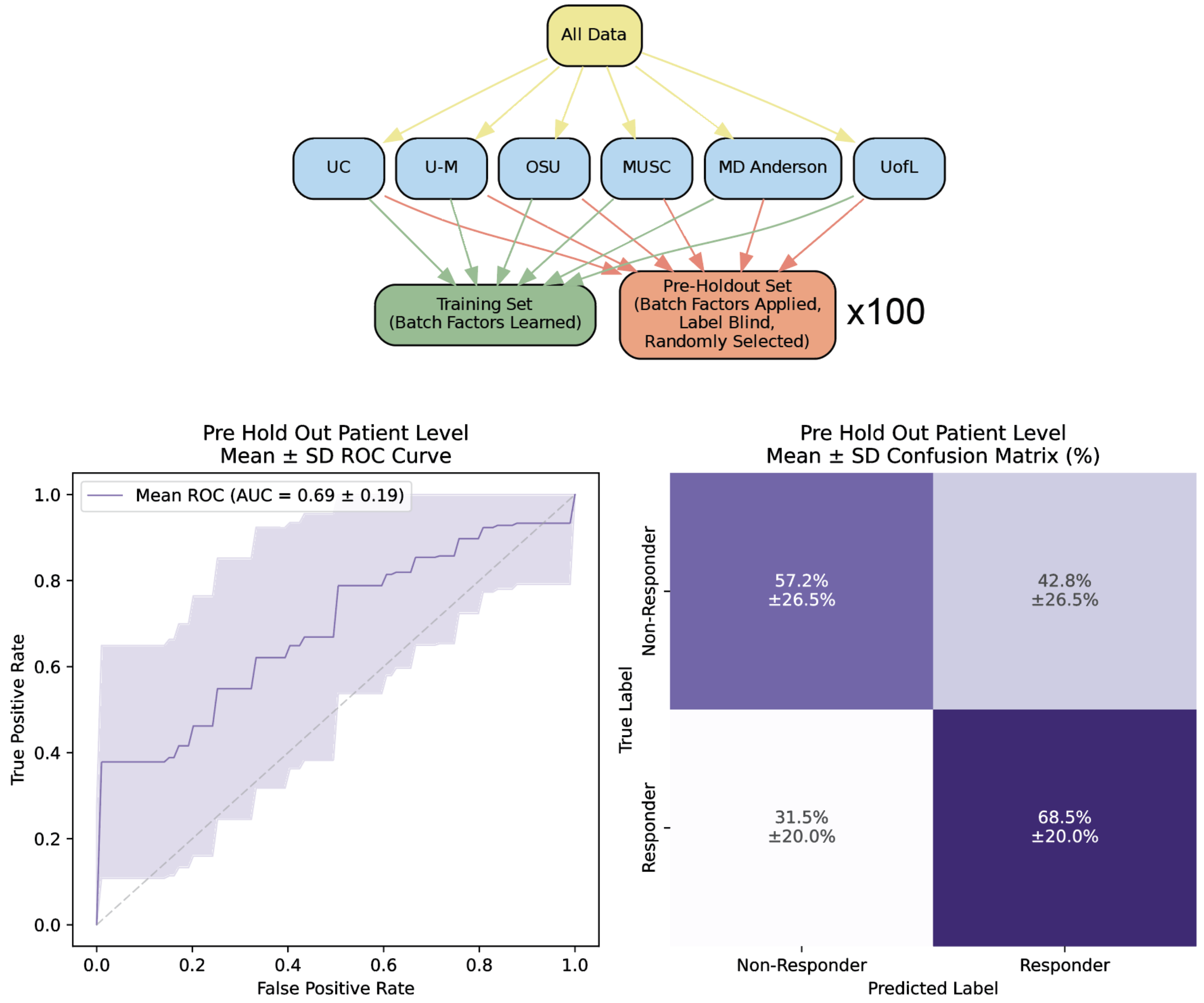


**Supplementary Figure 14. Patient-level performance of rMDS model across clinical timepoints in the pre hold-out evaluation strategy.** Models were trained using random 10-patient hold-outs repeated 100 times. ROC AUC values and their standard deviation are indicated.

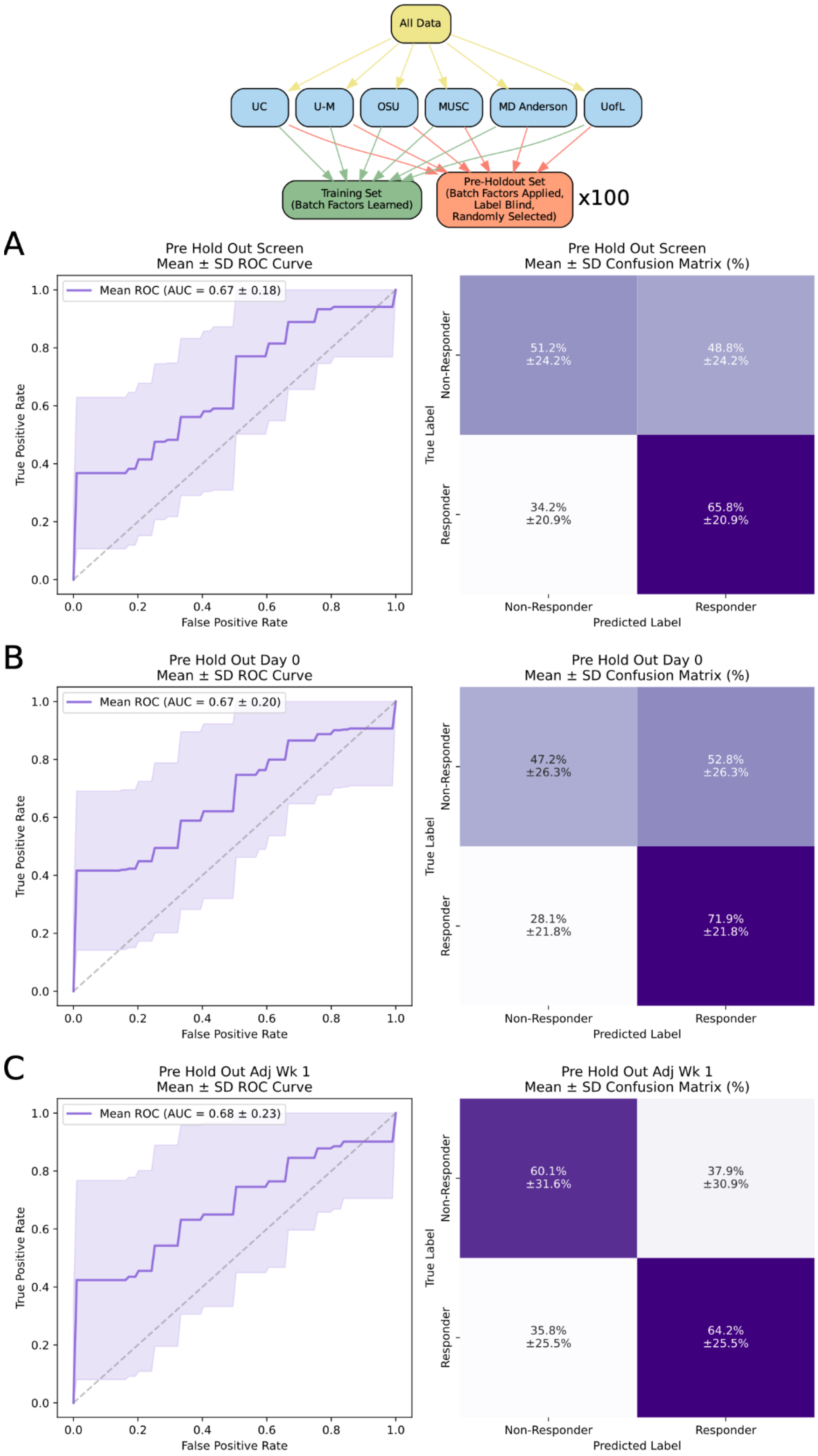


**Supplementary Figure 15. Sample-level performance of rMDS model across clinical timepoints in the pre hold-out evaluation strategy.** Receiver operating characteristic (ROC) curves and confusion matrices for the rMDS model evaluated on random 10-patient hold-outs repeated 100 times. Performance is shown overall and stratified by plasma collection timepoints: **(A)** Screen, **(B)** Day 0, and **(C)** Adjuvant Week 1. ROC AUC values and their standard deviation are indicated.

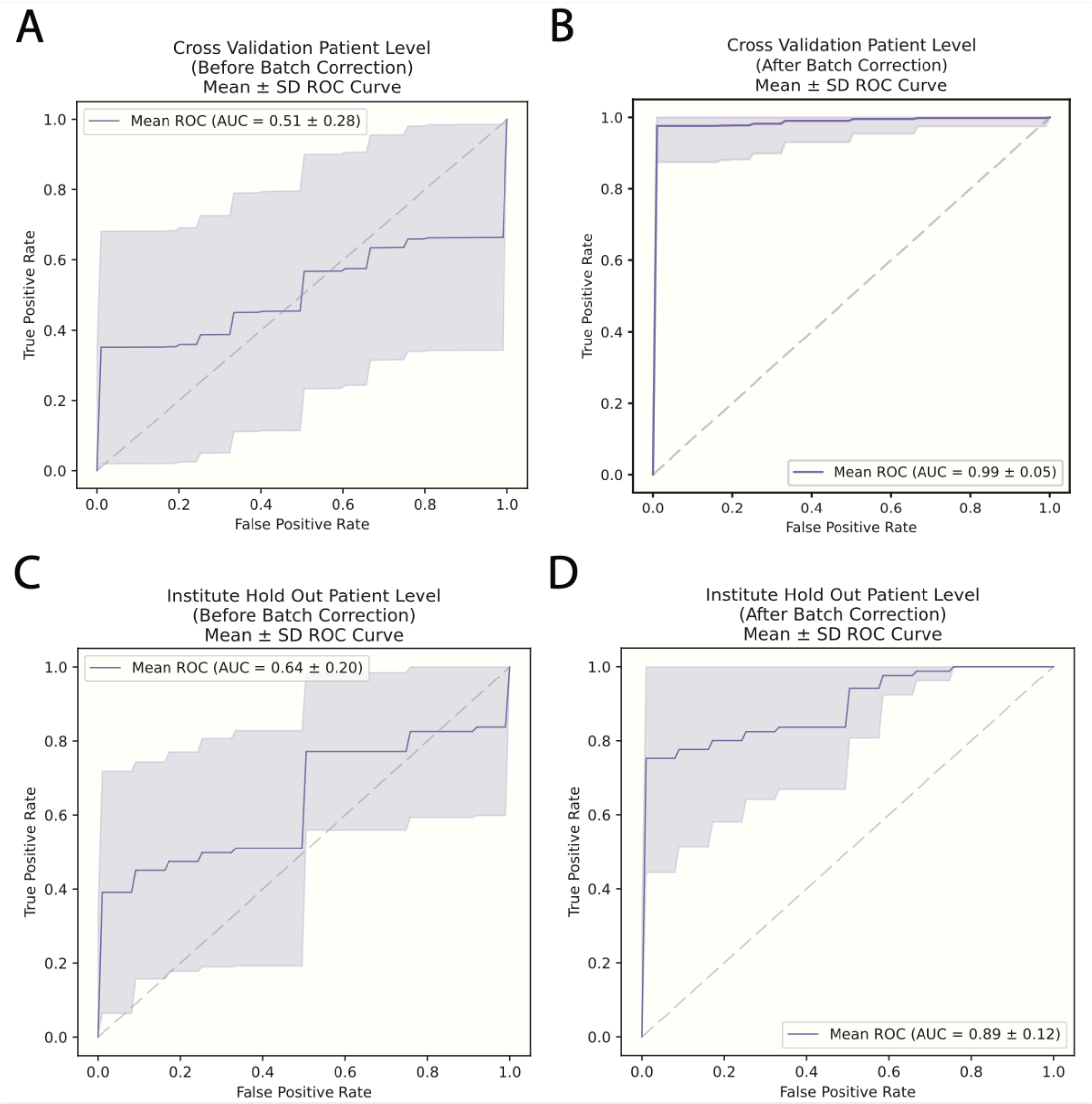


**Supplementary Figure 16. Predictive performance of rMDS before and after batch-correction of technical factors.** (A) Cross-validation evaluation strategy without batch correction and (B) after batch correction. Models were trained using a 10-fold patient-level cross-validation repeated 100 times. (C) Institute hold-out evaluation strategy without batch correction and (D) after batch correction. For each iteration, one institute was held out entirely for testing while the model was trained on the remaining institutions. The ROC curves (mean ± standard deviation) are shown.

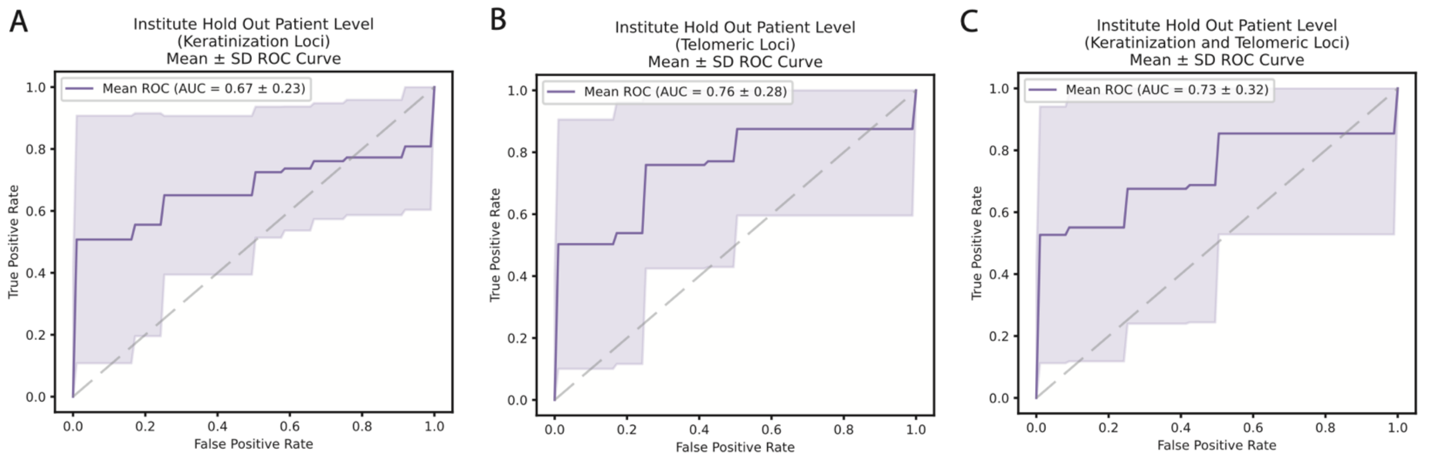


**Supplementary Figure 17. Predictive performance of rMDS using keratinization and telomeric loci.** (A) Institute hold-out evaluation strategy without keratinization loci and (B) telomeric loci and (C) both keratinization and telomeric loci. For each iteration, one institute was held out entirely for testing while the model was trained on the remaining institutions. The average ROC curve and confusion matrix (mean ± standard deviation) are shown.

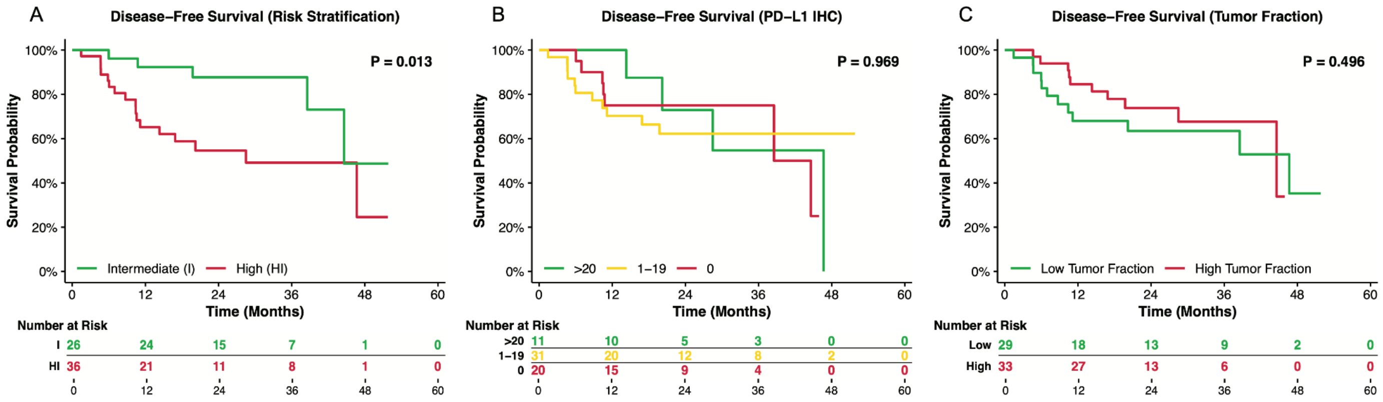


**Supplementary Figure 18. Kaplan-Meier survival analysis of disease-free survival stratified by cfDNA-derived and clinical biomarkers in HNSCC patients treated with pembrolizumab. (A)** Disease-free survival stratified by pathological risk (Intermediate, High). **(B)** Disease-free survival stratified by PD-L1 CPS (0, 1–19, >20). **(C)** Disease-free survival stratified by tumor fraction (Low vs High). Global log-rank p-values and risk tables are shown for each comparison.

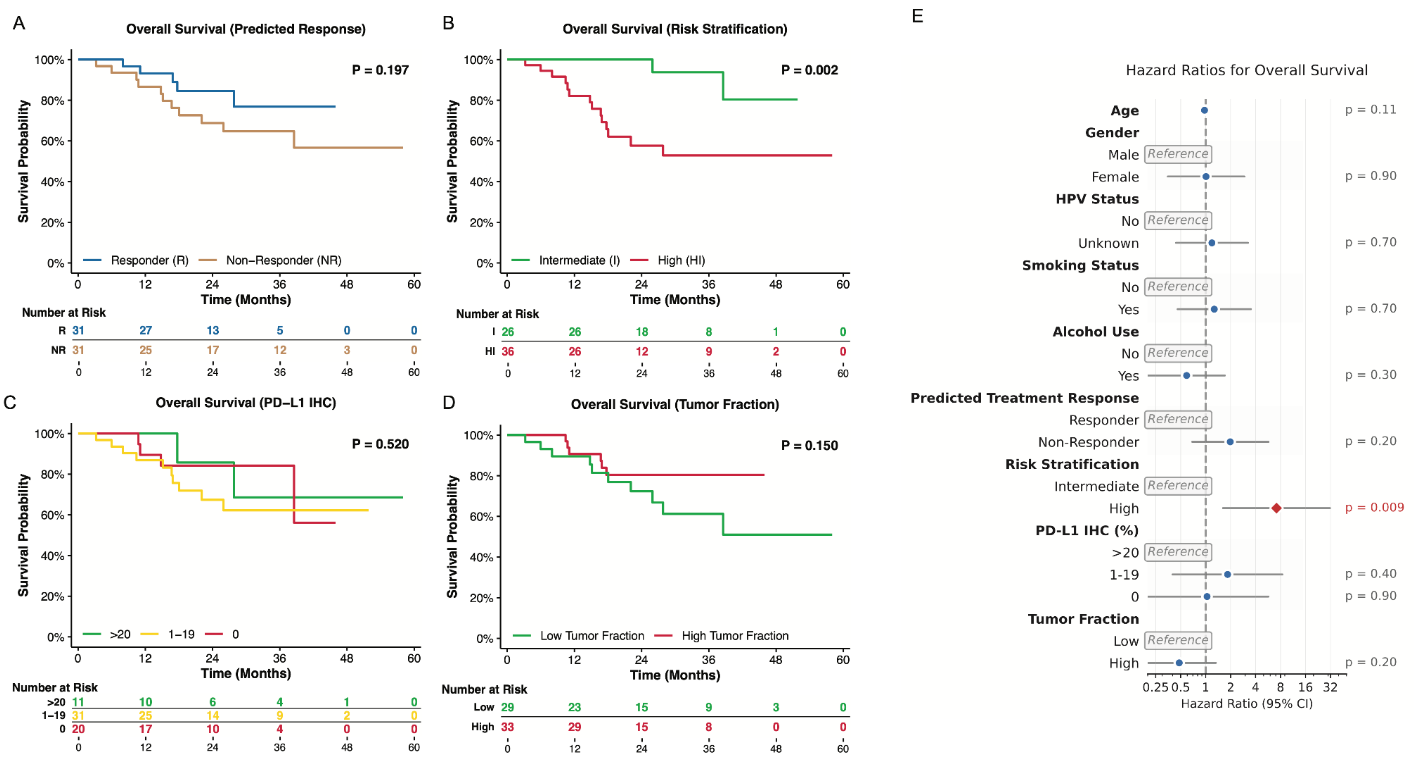


**Supplementary Figure 19. Kaplan-Meier survival analysis of overall survival stratified by cfDNA-derived and clinical biomarkers in HNSCC patients treated with pembrolizumab. (A)** Overall survival stratified by predicted response (Responder, Non-Responder) using a cfDNA rMDS-based model with single-institute hold-out evaluation strategy. **(B)** Overall survival stratified by pathological risk (Intermediate, High). **(C)** Overall survival stratified by PD-L1 CPS (0, 1–19, >20). **(D)** Overall survival stratified by tumor fraction (Low vs High). Global log-rank p-values and risk tables are shown for each comparison. **(E)** Hazard ratios for overall survival across clinical and demographic covariates (p-values indicated).

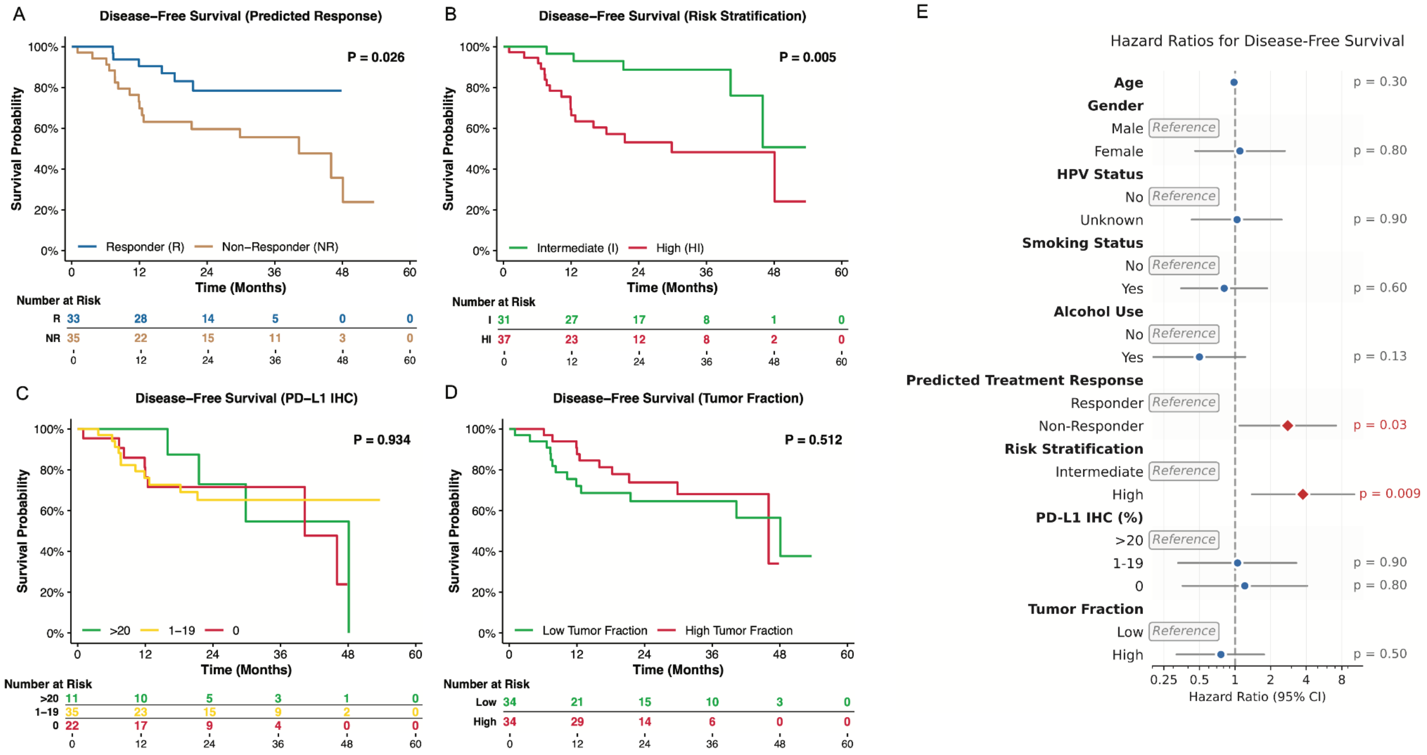


**Supplementary Figure 20. Kaplan-Meier survival analysis of disease-free survival based on time from neoadjuvant pembrolizumab stratified by cfDNA-derived and clinical biomarkers in HNSCC patients.** **(A)** Disease-free survival stratified by predicted response (Responder, Non-Responder) using a cfDNA rMDS-based model with single-institute hold-out evaluation strategy. **(B)** Disease-free survival stratified by pathological risk (Intermediate, High). **(C)** Disease-free survival stratified by PD-L1 CPS (0, 1–19, >20). **(D)** Disease-free survival stratified by tumor fraction (Low vs High). Global log-rank p-values and risk tables are shown for each comparison. **(E)** Hazard ratios for disease-free survival across clinical and demographic covariates (p-values indicated).

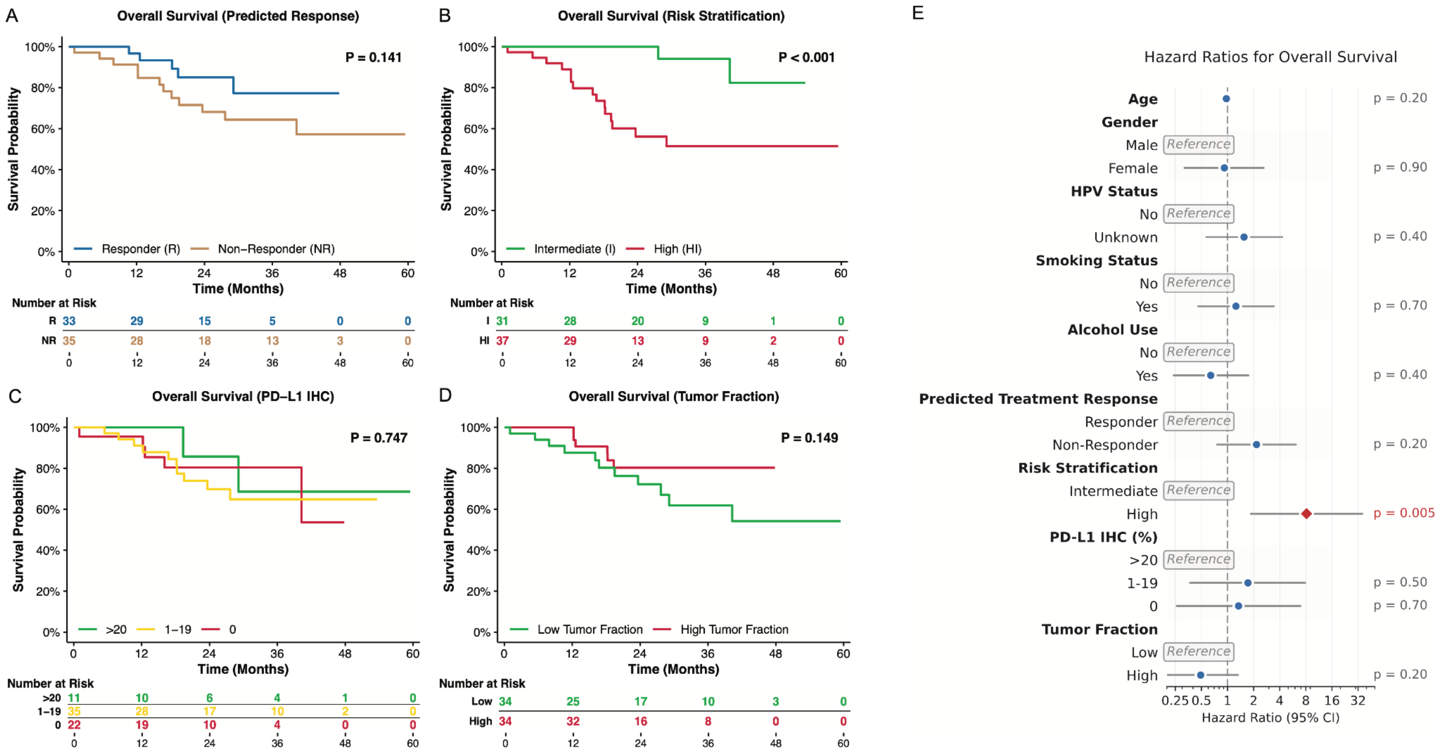

**Supplementary Figure 21. Kaplan-Meier survival analysis of overall survival based on time from neoadjuvant pembrolizumab stratified by cfDNA-derived and clinical biomarkers in HNSCC patients. (A)** Overall survival stratified by predicted response (Responder, Non-Responder) using a cfDNA rMDS-based model with single-institute hold-out evaluation strategy. **(B)** Overall survival stratified by pathological risk (Intermediate, High). **(C)** Overall survival stratified by PD-L1 CPS (0, 1–19, >20). **(D)** Overall survival stratified by tumor fraction (Low vs High). Global log-rank p-values and risk tables are shown for each comparison. **(E)** Hazard ratios for overall survival across clinical and demographic covariates (p-values indicated).

### Supplementary Tables

- **Supplementary Table 1:** Clinical metadata for all patients and plasma samples.
- **Supplementary Table 2:** cfDNA whole-genome sequencing (WGS) quality metrics.
- **Supplementary Table 3:** Silhouette scores for rMDS discrimination across various fragmentation features.
- **Supplementary Table 4:** Differentially enriched rMDS regions across all samples, separated by clusters.
- **Supplementary Table 5:** Gene ontology and pathway enrichment results for differential rMDS regions.
- **Supplementary Table 6:** Model performance metrics for cross-validation evaluation, institute-holdout evaluation, and pre-holdout evaluation.
- **Supplementary Table 7:** Hazard ratios for disease-free survival and overall survival.
